# No such thing as a free-rider? Understanding multicountry drivers of childhood and adult vaccination

**DOI:** 10.1101/2020.12.07.20245118

**Authors:** Frederik Verelst, Roselinde Kessels, Lander Willem, Philippe Beutels

## Abstract

**Background:** Increased vaccine hesitancy and refusal negatively affects vaccine uptake leading to vaccine preventable disease reemergence. We aimed to quantify the relative importance of characteristics people consider when making vaccine decisions for themselves, or for their child, with specific attention for underlying motives arising from context, such as required effort (accessibility) and opportunism (free riding on herd immunity).

**Methods:** We documented attitudes towards vaccination and performed a discrete choice experiment in 4802 respondents in The United Kingdom, France and Belgium eliciting preferences for six attributes: (1) vaccine effectiveness, (2) vaccine preventable disease burden, (3) vaccine accessibility in terms of co-payment, vaccinator and administrative requirements, (4) frequency of mild vaccine-related side-effects, (5) vaccination coverage in the country’s population and (6) local vaccination coverage in personal networks. We distinguished adults deciding on vaccination for themselves (‘oneself’ group) from parents deciding for their youngest child (‘child’ group).

**Results:** While all six attributes were found to be significant, vaccine effectiveness and accessibility stand out in all (sub)samples, followed by vaccine preventable disease burden. We confirmed that people attach more value to severity of disease compared to its frequency and discovered that peer influence dominates free-rider motives, especially for the vaccination of children.

**Conclusions:** These behavioral data are insightful for policy and are essential to parameterize dynamic vaccination behavior in simulation models. In contrast to what most game theoretical models assume, social norms dominate free-rider incentives. Therefore policy-makers and healthcare workers should actively communicate on high vaccination coverage, and draw attention to the effectiveness of vaccines, while optimizing their practical accessibility.

## Background

Vaccination remains a cornerstone of global public health, preventing about 2 to 3 million deaths each year [1]. However, its success is currently undermined by growing vaccine hesitancy and refusal. Sentiments underpinning this have multi-faceted origins, not least distorted perceptions of severe vaccine side-effects, much of which can be traced back to fraudulent research linking measles-mumps-rubella (MMR) vaccination with autism [2, 3], and misconceptions about the use of adjuvants in vaccines [4]. Others include doubts about vaccine effectiveness [3, 5] and about our immune system’s coping with the rising number of recommended vaccine antigens [3, 6]. More extreme attitudes are based on government and vaccine industry conspiracy theories [3], religious beliefs (e.g. Protestantism in the Dutch Bible Belt [7]) and “back to nature” motives (i.e. preferring immunity acquired by natural infection to vaccine-induced immunity, under the belief that “divine or natural” risks are smaller and/or more “just” than those imposed by human interventions) [3].

Even though vaccine controversies are not new [8, 9], the internet and a variety of social media have amplified the spread of misinformation and allowed the establishment of new online anti-vaccine communities [10]. According to a 2018 Gallup poll [11], only 40% and 59% of Eastern and Western Europeans, respectively, believe vaccines are safe. In Northern Europe and Northern America, these figures are higher at 73% and 72%, respectively [11].

As a result of these misperceptions, plunging vaccination rates and immunity levels have been observed in recent years. Notably so for measles, which is a highly virulent pathogen for which a safe and effective vaccine was already approved by the Food and Drug Administration (FDA) in 1971 [12]. Indeed, the European Centre for Disease Prevention and Control (ECDC) recently reported the existence of a large pool of people in the EU that are susceptible to measles due to low historical and current vaccination coverage. Only 4 countries achieved two dose measles vaccination coverage of at least 95% in 2017, compared to 14 countries in 2007. Unsurprisingly, measles resurgence has recently been observed, with 44,074 cases in 30 EU member states between 2016 and March 2019 [13]. The same trend has been observed in the US, with 704 cases reported in the first four months of 2019 (even though the US declared elimination of endemic transmission in 2000) [14, 15].

Mathematical and economic models have proven valuable to simulate and evaluate the impact of prevention measures on the spread, burden and economics of infectious diseases. These models inform and guide policy-makers to prepare for and respond to (re)emerging infectious diseases, particularly when sufficient information from controlled experiments is lacking. However, because of the reasons previously touched upon, the impact of prevention measures and other policy interventions are subject to hosts’ compliance and demand. In response, behavioral change models have been developed to incorporate dynamic behavior (i.e. the demand side of prevention measures) into models for infectious disease transmission. As a result of circulating controversies and -usually positive-externalities, vaccination models have become particularly interesting to take dynamic behavior into account. Indeed, vaccination usually results in positive externalities, often referred to as ‘herd immunity’: successfully vaccinated individuals do not (or hardly) transmit the pathogen to others. As such the marginal utility of vaccination decreases (non-linearly) as coverage increases, and endemic transmission can often be halted without vaccinating the whole population, a phenomenon which is crucial for vulnerable individuals who cannot receive vaccination due to age or medical reasons (e.g. too young or immuno-compromised). Where positive externalities exist, game theory applies. Hence, models have been developed in which rational-behaving individuals are assumed to free-ride on ‘herd immunity’, and therefore increasingly refuse vaccination when they perceive more members of the population to be immunized. However, the majority of behavioral change models in the published literature remains purely theoretical, lacking parameterization with empirical data and a validation process [16, 17]. Consequently, data for parameterization of behavioral change models are highly desirable to construct improved models mimicking realistic vaccination behavior. This is generally recognized as one of the challenges for behavioural change models [18].

Discrete choice experiments (DCEs) have proven successful to elicit preferences and quantify the decision-making process with respect to vaccine characteristics in multiple studies [19–26]. Moreover, they are well established as an instrument in health economic research in general [27]. A DCE is a quantitative surveying technique in which respondents make a choice between two or more hypothetical profiles in consecutive choice sets. Profiles are represented by attributes with (partially) differing attribute levels [28]. In previous DCEs, vaccines were described using attributes such as vaccine effectiveness [19, 20, 22–25, 29], vaccine-related side-effects (VRSE) [19–24, 29] or in terms of vaccine price (whether or not including costs of vaccine administration) [19, 21–24, 26]. A recent study found that DCEs correctly predicted influenza vaccination choices on an aggregate level when taking scale and preference heterogeneities into account [30].

In this paper, we report on the findings of a DCE quantifying individual preferences for vaccination attributes in Belgium, the United Kingdom (UK) and France. We present these new results together with those of two separately reported DCEs using an identical design, conducted in South Africa and The Netherlands [24, 31]. We aim to: 1) generate and communicate behavioral data with respect to vaccines in order to move from theory to data-driven behavioral change models in infectious disease epidemiology, 2) assess to what extent individual vaccination decisions are driven by social norms or peer pressure as opposed to free-riding motives, 3) identify the vaccine characteristics society values most, and 4) accommodate policy-makers and health care professionals to select focal points in their communication to hesitant individuals.

## Methods

We conducted a survey in France, the UK (both early December 2018) and Belgium (May 2019). We selected these countries for a number of reasons. First of all, no DCE had yet been performed for a general, unnamed vaccine, distinguishing between adults and children in any of these countries. Also, we were interested in between-country differences comparing different backgrounds, cultures and more specifically, a different history with respect to vaccination. France was included in this study because it has been experiencing a lot of vaccine resistance: one in three French inhabitants now believes vaccines are unsafe, which is the highest fraction in the world [11]. More specifically, there is a lot of vaccine resistance in France originating from safety concerns regarding the pandemic A/H1N1 flu vaccine with spillovers to other vaccines (e.g. MMR vaccine) [11, 32]. As a result, the French government expanded the number of compulsory vaccines from 3 to 11 in 2018 [11]. The UK was included because it has a history of vaccine scares with documented impact on vaccine coverage for the whole cell pertussis vaccine in the 1970s and 1980s [33] and MMR vaccine in the 2000s [2]. We also included Belgium, a country with a more neutral vaccination history, achieving generally high and stable vaccine coverage in young children [34]. However, regional disparities have been observed to widen [35], and one in five Belgian citizens believe vaccines are unsafe [11]. In order to facilitate broader between-country comparisons, we report our results alongside those of two more studies using an identical design in South Africa and The Netherlands, conducted in December 2017 and June-July 2018, respectively, and published in detail elsewhere [24, 31].

Given that the design of the questionnaire was explicitly developed for cross-country comparisons, the majority of the survey questions and the entire DCE design were kept the same as in South Africa and The Netherlands, which in turn were based on a study in Flanders in 2017 [23]. The survey questionnaire is provided as a supplement to the paper by Hoogink et al. [31] and as a supplement to this paper (additional file 2). We adapted the survey questions to reflect country-specific characteristics based on inputs from local experts, for example with respect to the educational system and the organization of the national immunization schedule. As such, we ended up with four versions of the survey for the UK, France, French speaking Belgium, and Dutch speaking Belgium. The multi-country study protocol as well as the amendments to the original questionnaires (Reference number: 15/2/12) were reviewed and accepted by the Ethical Committee of the Antwerp University Hospital (UZA, Belgium). Given that this study was non-interventional, and carried out in a general population (adults only) with informed consent, duplicate ethical approval at a local committee was not deemed necessary by our IRB, and we therefore did not re-apply for it with a local committee. We tested each survey version in a soft launch in which we asked about 10% of the target sample to fill out the survey and evaluate the comprehensibility of the questions. Afterwards, we launched the survey in the sample population. The survey consisted of five sections: 1) background questions probing for age, gender, marital status, occupation, smoking behavior, etc., 2) 21 attitudinal questions on vaccines where responses were recorded on a five-point Likert scale, ranging from completely agree to completely disagree, 3) a DCE with 10 choice sets based on Verelst et al. [23], including an introduction text with instructions and a sample choice set to familiarize the respondents with the DCE, 4) four questions probing for relative risk perceptions based on a survey by Bults et al. [36], and 5) a health literacy test with three questions from Chew et al. [37]. Based on their background characteristics, we allocated respondents to two distinct surveys: a ‘oneself’ group (without allocation restrictions) and a ‘youngest child’ group (only for respondents having at least one child below the age of 18 years), the former filling out the survey with respect to vaccination decisions for themselves, the latter doing so for their youngest child. We opted for a sample size of about 1500 respondents per country, based on previous DCEs with the same design [23, 24, 31]. We gradually built each sample to better match the sample demographics to the population demographics, and thus to obtain a more representative sample. We recruited respondents from an online consumer panel applying an efficient participant allocation algorithm. In total, 9339 respondents started the survey, 4802 of them completed the survey, 1213 chose not to complete it, 59 did not meet the inclusion criteria (e.g., <18 years old), 119 were identified as ‘speeders’ (who filled out the survey much faster than a reference time) and/or ‘straight-liners’ (who filled out the same for each question), and 3146 were halted after the first part of the survey with background questions when pre-defined sample quota were reached. We incentivized participation through credit rewards, transferable into coupons and gift vouchers. Only one member per household could participate in the study. Country and group level sample characteristics are displayed in Table 1.

**Table 1.** Sample characteristics and national statistics for Belgium, France & The UK.

| Characteristic | Belgium |  | United Kingdom |  | France |  |
| --- | --- | --- | --- | --- | --- | --- |
|  | Sample (%) | Population (%) | Sample (%) | Population (%) | Sample (%) | Population (%) |
| **Gender** |  |  |  |  |  |  |
| Male | 50.2 | 49.1 | 45.8 | 49.1 | 40.7 | 48.4 |
| Female | 49.8 | 50.9 | 54.2 | 50.9 | 59.3 | 51.6 |
| **Age group (\*)** |  |  |  |  |  |  |
| 18-34 | 26.6 | 22.9 | 24.8 | 24.9 | 27.1 | 23.5 |
| 35-49 | 26.4 | 34.7 | 36.4 | 34.7 | 39.6 | 34.2 |
| 50-65 | 25.4 | 24.0 | 26.9 | 22.7 | 24.6 | 24.2 |
| 66-85 | 21.6 | 18.4 | 12.0 | 17.7 | 8.8 | 18.1 |
| **Educational attainment** |  |  |  |  |  |  |
| Primary education (ISCED 1) or lower | 8.3 | 14.6 | <1 | <1 | 1.1 | 17.5 |
| Secondary education (ISCED 2+3) | 55.6 | 49.6 | 58.4 | 70.0 | 72.2 | 58.3 |
| Post-secondary or (post-)university education (ISCED 4 or higher) | 33.7 | 26.3 | 39.3 | 30.0 | 25.8 | 24.2 |
| Other | 2.4 | 9.5 | 1.8 | <1 | <1 | <1 |
| **NUTS 1 region** |  |  |  |  |  |  |
| **Belgium** |  |  |  |  |  |  |
| Flanders | 57.4 | 57.5 |  |  |  |  |
| Walloon region | 30.2 | 32.2 |  |  |  |  |
| Brussels Capital Region | 12.3 | 10.3 |  |  |  |  |
| **UK** |  |  |  |  |  |  |
| North East |  |  | 5.4 | 4.1 |  |  |
| North West |  |  | 10.0 | 11.2 |  |  |
| Yorkshire and the Humber |  |  | 8.6 | 8.4 |  |  |
| East Midlands |  |  | 6.6 | 7.2 |  |  |
| West Midlands |  |  | 9.7 | 8.9 |  |  |
| East of England |  |  | 8.9 | 9.3 |  |  |
| London |  |  | 11.2 | 12.9 |  |  |
| South East |  |  | 12.6 | 13.7 |  |  |
| South West |  |  | 8.3 | 8.4 |  |  |
| Wales |  |  | 6.3 | 4.8 |  |  |
| Scotland |  |  | 8.6 | 8.4 |  |  |
| Northern Ireland |  |  | 3.9 | 2.9 |  |  |
| **France** |  |  |  |  |  |  |
| Région parisienne |  |  |  |  | 16.3 | 18.3 |
| Bassin parisien |  |  |  |  | 22.1 | 16.6 |
| Nord |  |  |  |  | 5.6 | 6.2 |
| Est |  |  |  |  | 10.8 | 8.3 |
| Ouest |  |  |  |  | 12.9 | 13.2 |
| Sud-Ouest |  |  |  |  | 11.3 | 10.6 |
| Centre-Est |  |  |  |  | 10.0 | 11.8 |
| Méditerranée |  |  |  |  | 10.8 | 12.2 |
| Départements d'Outre Mer |  |  |  |  | 0.2 | 2.9 |
| **Sample size** | **N=1602** |  | **N=1600** |  | **N=1600** |  |
| ‘Oneself’ | N=1001 |  | N=850 |  | N=850 |  |
| ‘Youngest child’ | N=601 |  | N=750 |  | N=750 |  |

\*age groups from the survey are compared to age groups [15-29], [30-49], [50-64] and [65-84] as reported in the 2011 census database [40].

The DCE was characterized by a Bayesian D-efficient design [28] of 50 choice sets with 2 profiles described by 6 attributes with 3 varying and 3 constant levels and optimized for the precise estimation of all main effects as well as all two-way interactions between any of the six attributes and ‘vaccine effectiveness’, ‘VRSE’ and ‘accessibility’. We divided the design into five surveys of 10 choice sets that we distributed evenly between all participants. We selected the attributes and attribute levels through a literature study, a focus group study and a pilot study in Flanders, the details of which are published in Verelst et al. [23]. We revised the description of VRSE by specifying the severity of side-effects, keeping severe side-effects to be ‘highly unlikely’ in the two profiles, and only varying the frequency of mild VRSE. This contrasts with the original design in Verelst et al. [23], where we left the severity of side-effects unspecified, but is the same as in Verelst et al. [24] and Hoogink et al. [31]. We opted for this strategy since it prevents the participant from imagining levels regarding VRSE severity and it mimics real-life VRSE, because vaccines with common severe side-effects should not be licensed. We included population and local coverage as attributes to assess the magnitude of free-riding behavior in the populations under study. Negative utility values for higher coverage levels confirm free-riding behavior, as opposed to positive utility values, in which case peer influence and social norms dominate. All attributes and attribute levels are shown in Table 2.

**Table 2.**
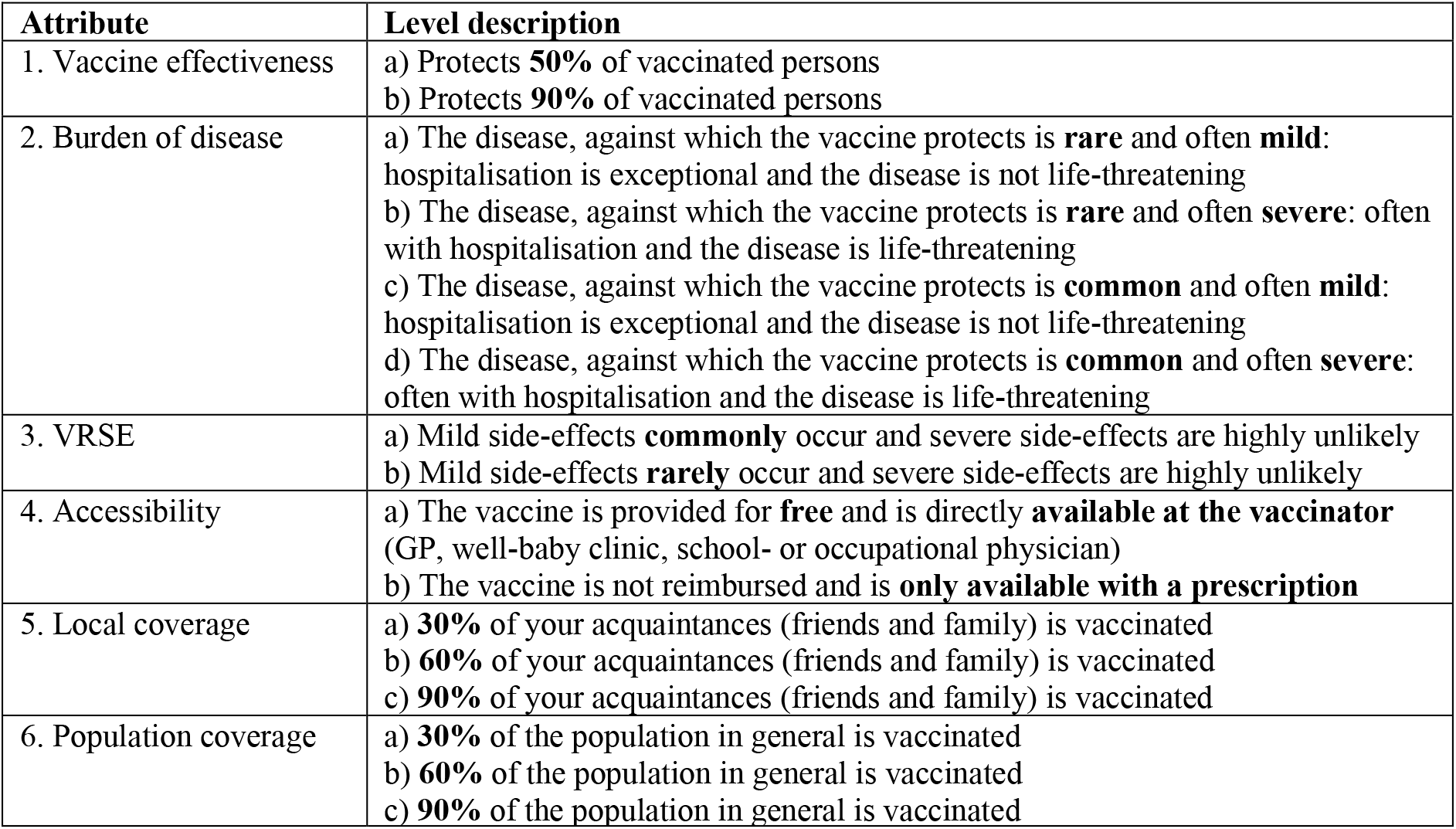
DCE attributes and levels.

| Attribute | Level description |
| --- | --- |
| 1. Vaccine effectiveness | a) Protects **50%** of vaccinated persons<br>b) Protects **90%** of vaccinated persons |
| 2. Burden of disease | a) The disease, against which the vaccine protects is **rare** and often **mild**: hospitalisation is exceptional and the disease is not life-threatening<br>b) The disease, against which the vaccine protects is **rare** and often **severe**: often with hospitalisation and the disease is life-threatening<br>c) The disease, against which the vaccine protects is **common** and often **mild**: hospitalisation is exceptional and the disease is not life-threatening<br>d) The disease, against which the vaccine protects is **common** and often **severe**: often with hospitalisation and the disease is life-threatening |
| 3. VRSE | a) Mild side-effects **commonly** occur and severe side-effects are highly unlikely<br>b) Mild side-effects **rarely** occur and severe side-effects are highly unlikely |
| 4. Accessibility | a) The vaccine is provided for **free** and is directly **available at the vaccinator** (GP, well-baby clinic, school- or occupational physician)<br>b) The vaccine is not reimbursed and is **only available with a prescription** |
| 5. Local coverage | a) **30%** of your acquaintances (friends and family) is vaccinated<br>b) **60%** of your acquaintances (friends and family) is vaccinated<br>c) **90%** of your acquaintances (friends and family) is vaccinated |
| 6. Population coverage | a) **30%** of the population in general is vaccinated<br>b) **60%** of the population in general is vaccinated<br>c) **90%** of the population in general is vaccinated |

We analysed the DCE using the JMP Pro 14 Choice Platform [38] and applied a Panel Mixed Logit (PML) modeling approach with 10,000 Bayesian iterations, with the last 5,000 used for estimation. We distinguished between models estimating the attribute effects - allowing for model comparison between study populations - and models including interaction effects between the attributes and respondent covariates - allowing for identifiable preference heterogeneity within study populations. In the latter, we systematically estimated covariate interactions one-by-one, keeping record of all the statistically significant model terms including the main effects. Afterwards, we estimated a joint model combining all main effects and individually significant interactions. We dropped insignificant interactions in an iterative process until we reached a model with the most important covariates. We ranked the significant model terms by importance using the normalized LogWorth statistic, i.e. -log_10_(p-value of the LR-test), where the LR-test is short for the likelihood ratio test for significance of a given model term. We used RStudio [39] for cleaning the raw survey data and creating the bar charts.

## Results

We managed to retrieve a quasi-representative sample of about 1600 survey respondents in each country, as shown in Table 1. Women are slightly overrepresented in the samples from the UK and France. We found a representative population with respect to age to be incompatible with having at least 750 respondents with children below the age of 18. Moreover, concerning educational attainment, the samples are also somewhat biased towards the higher educated, especially so in France. This is likely because older French respondents, who tend to be lower educated, are underrepresented in our sample. Note however, that the youngest age groups are by definition lower educated since the census data also include school-age teenagers (15-18 years). We investigated the impact of mismatching sample characteristics by estimating covariate interactions between the attributes and gender, educational attainment and region, and found none of them to significantly influence our findings. Significant covariate interactions with respondents’ age group are included and reported in Appendix A.

Vaccine attitudes tended to be positive in general as represented in Figures 1 and 2 for a selection of general vaccine statements. We observed French respondents in the ‘adult’ group to be relatively neutral towards the statements “The people who are important to me think that I must get vaccinated” and “I have confidence in the information about vaccinations that I receive from the Government”. These sentiments appeared to be more negative in the ‘adult’ group than for the ‘child group’. In contrast, the respondents from the UK were in general more agreeing on this selection of statements. The median UK respondent strongly agreed with the statements “I think that getting vaccinated against infectious diseases is wise” and “I think that getting vaccinated against infectious diseases is important” in the ‘adult’ group, whereas in the ‘child’ group the median UK respondent strongly agreed with the statements “The diseases that are vaccinated against can be very serious” and “I think that vaccinating my child according to the National Vaccination Program is important”. Other ‘child’ group samples, on average, agree on all statements, though there is a lot of variability within the samples. Attitudes from Belgian and Dutch respondents were usually found in between the UK and the French sample means. Details on all 21 attitudinal questions are presented in Appendix B.

**Figure 1.**
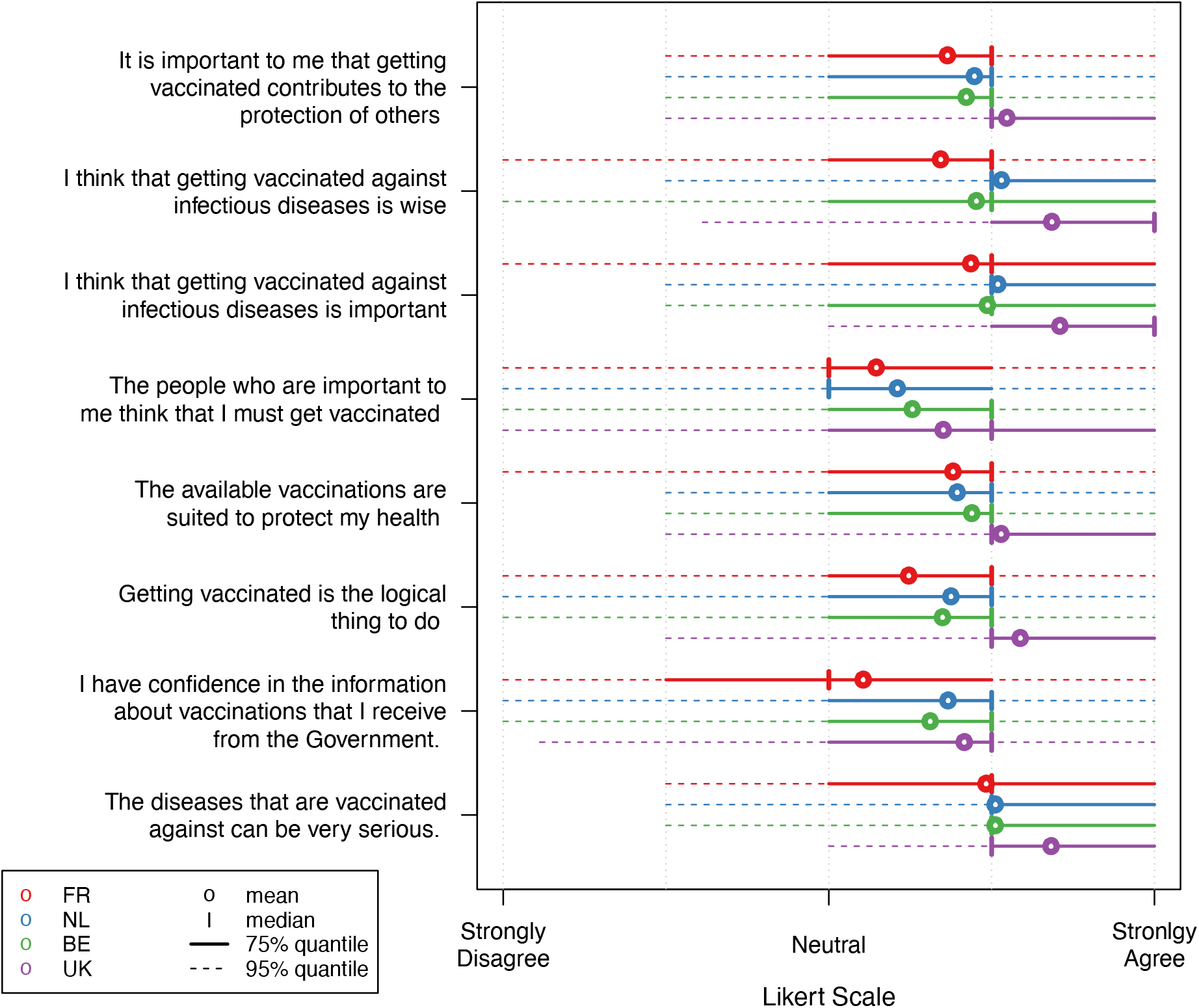
Likert scale responses for a selection of vaccination attitude statements in the ‘adult’ group.

**Figure 2.**
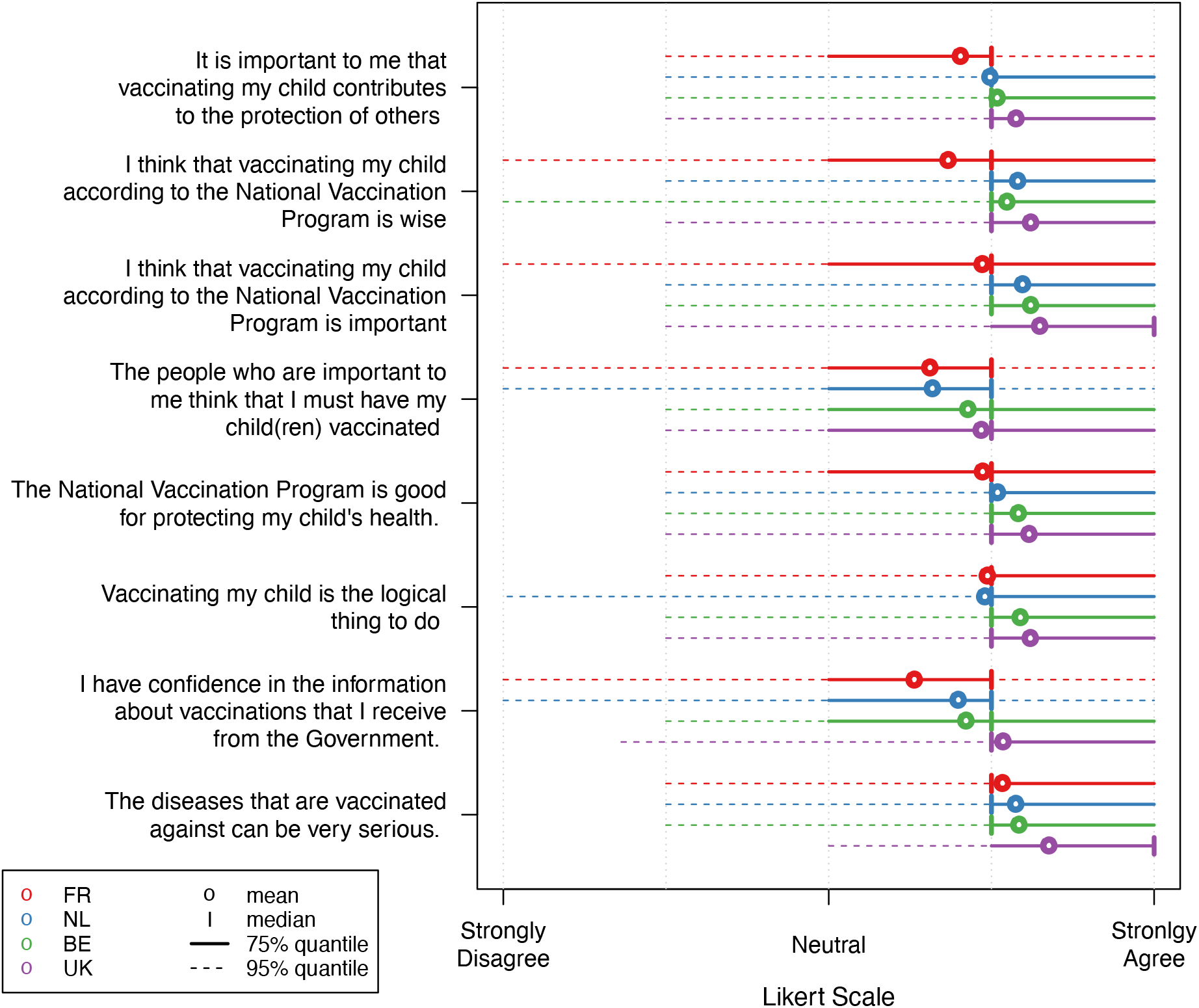
Likert scale responses for a selection of vaccination attitude statements in the ‘child’ group.

All six attributes were found to be statistically significant in all five countries. The normalized LogWorth values represent the relative importance of the attributes in each country and subgroup, and are visualized in Figure 3. Two attributes stand out: vaccine effectiveness and accessibility. Vaccine effectiveness is the key characteristic for all survey respondents in the UK and South Africa, but also for the ‘child’ group in The Netherlands. For the Belgian population as well as the French ‘oneself’ group, accessibility was found to be most important. The French ‘child’ group attached most importance to burden of disease, whereas this was considered much less important by the same subpopulation in the UK. We found local coverage and mild VRSE to be also statistically significant but of limited importance in most study samples, with a relative importance of 30% or less. Population coverage was found to have more influence, especially so in the case of ‘child’ models, with the Netherlands being an exception. Note that among all five countries mild VRSE had the highest impact in vaccine decision-making in France and Belgium.

**Figure 3.**
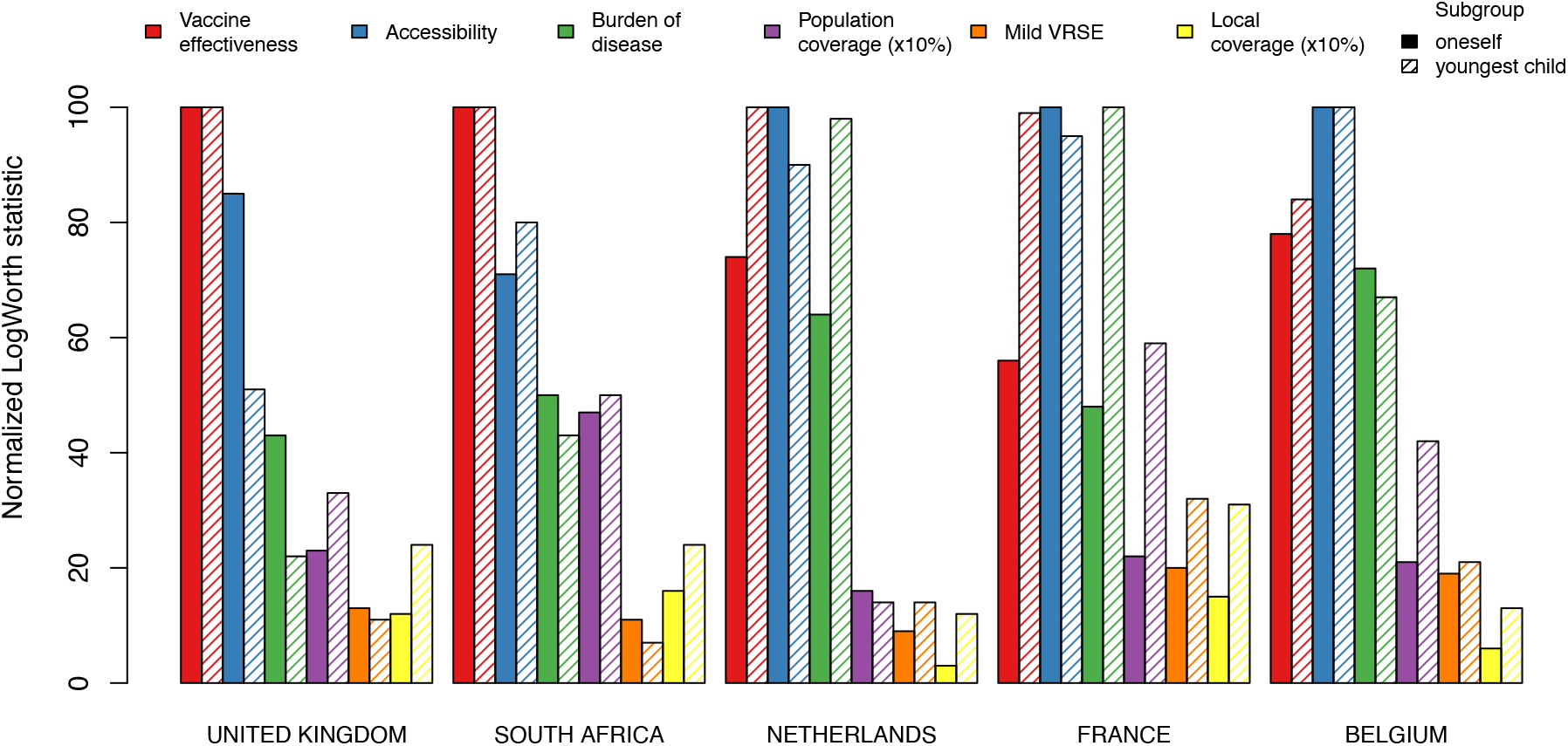
Importance of all statistically significant (p-value <0.05) main effects relative to the most important attribute in each study sample.

For both population and local coverage, estimates were found to be positive for all subsamples in all study countries (see Tables 3 to 5). Hence, respondents were more inclined to choose a vaccine if it already had a high coverage in their network of contacts and in the population at large. For example, for the ‘child’ group in France, a 10% increase in the population’s vaccination coverage increases vaccine utility by 0.108 on average (see Table 5).

**Table 3.** Panel mixed logit model estimates (means and standard deviations) and significances of the attribute effects obtained from likelihood ratio (LR) tests. **Belgium**

| Term | Mean estimate (std dev;<br>subject std dev) | LR Chi-Square | DF | P-value |
| --- | --- | --- | --- | --- |
| **‘Oneself’ model** |  |  |  |  |
| **Accessibility**<br>Co-payment & prescription<br>Free & accessible | -0.403 (0.020; 0.334)<br>0.403 (0.020; 0.316) | 326.606 | 1 | <0.0001 |
| **Vaccine effectiveness**<br>50%<br>90% | -0.465 (0.024; 0.244)<br>0.465 (0.025; 0.210) | 252.171 | 1 | <0.0001 |
| **Burden of disease**<br>Rare & mild<br>Common & mild<br>Rare & severe<br>Common & severe | -0.436 (0.045; 0.615)<br>-0.481 (0.047; 0.361)<br>0.324 (0.037; 0.128)<br>0.593 (0.043; 0.174) | 243.682 | 3 | <0.0001 |
| **Population coverage (x10%)** | 0.081 (0.008; 0.099) | 65.749 | 1 | <0.0001 |
| **Mild VRSE**<br>Common<br>Rare | -0.184 (0.019; 0.098)<br>0.184 (0.020; 0.091) | 57.931 | 1 | <0.0001 |
| **Local coverage (x10%)** | 0.043 (0.008; 0.079) | 17.977 | 1 | <0.0001 |
| **‘Youngest child’ model** |  |  |  |  |
| **Accessibility**<br>Co-payment & prescription<br>Free & accessible | -0.472 (0.031; 0.346)<br>0.472 (0.029; 0.337) | 228.127 | 1 | <0.0001 |
| **Vaccine effectiveness**<br>50%<br>90% | -0.571 (0.037; 0.245)<br>0.571 (0.039; 0.226) | 191.508 | 1 | <0.0001 |
| **Burden of disease**<br>Rare & mild<br>Common & mild<br>Rare & severe<br>Common & severe | -0.370 (0.058; 0.463)<br>-0.613 (0.062; 0.418)<br>0.307 (0.056; 0.284)<br>0.676 (0.061; 0.348) | 161.860 | 3 | <0.0001 |
| **Population coverage (x10%)** | 0.128 (0.012; 0.126) | 93.449 | 1 | <0.0001 |
| **Mild VRSE**<br>Common<br>Rare | -0.234 (0.028; 0.137)<br>0.234 (0.031; 0.129) | 45.280 | 1 | <0.0001 |
| **Local coverage (x10%)** | 0.071 (0.013; 0.123) | 27.429 | 1 | <0.0001 |
| Note: Mean estimates corresponding to the last level of an attribute are calculated as minus the sum of the estimates for the other levels of the attribute. |  |  |  |  |

**Table 4.** Panel mixed logit model estimates (means and standard deviations) and significances of the attribute effects obtained from likelihood ratio (LR) tests. **United Kingdom**

| Term | Mean estimate (std dev;<br>subject std dev) | LR Chi-Square | DF | P-value |
| --- | --- | --- | --- | --- |
| **‘Oneself’ model** |  |  |  |  |
| **Vaccine effectiveness** |  |  |  |  |
| 50% | -0.683 (0.035; 0.275) | 425.353 | 1 | <0.0001 |
| 90% | 0.683 (0.031; 0.277) |  |  |  |
| **Accessibility** |  |  |  |  |
| Co-payment & prescription | -0.486 (0.023; 0.316) | 360.260 | 1 | <0.0001 |
| Free & accessible | 0.486 (0.027; 0.292) |  |  |  |
| **Burden of disease** |  |  |  |  |
| Rare & mild | -0.517 (0.049; 0.277) | 189.172 | 3 | <0.0001 |
| Common & mild | -0.351 (0.051; 0.430) |  |  |  |
| Rare & severe | 0.307 (0.045; 0.206) |  |  |  |
| Common & severe | 0.561 (0.051; 0.239) |  |  |  |
| **Population coverage (x10%)** | 0.096 (0.010; 0.118) | 94.330 | 1 | <0.0001 |
| **Mild VRSE** |  |  |  |  |
| Common | -0.180 (0.024; 0.124) | 50.290 | 1 | <0.0001 |
| Rare | 0.180 (0.025; 0.123) |  |  |  |
| **Local coverage (x10%)** | 0.080 (0.010; 0.078) | 47.291 | 1 | <0.0001 |
| **‘Youngest child’ model** |  |  |  |  |
| **Vaccine effectiveness** |  |  |  |  |
| 50% | -0.591 (0.033; 0.243) | 297.130 | 1 | <0.0001 |
| 90% | 0.591 (0.033; 0.246) |  |  |  |
| **Accessibility** |  |  |  |  |
| Co-payment & prescription | -0.309 (0.024; 0.233) | 149.559 | 1 | <0.0001 |
| Free & accessible | 0.309 (0.024; 0.218) |  |  |  |
| **Population coverage (x10%)** | 0.107 (0.009; 0.101) | 94.979 | 1 | <0.0001 |
| **Local coverage (x10%)** | 0.097 (0.010; 0.094) | 67.461 | 1 | <0.0001 |
| **Burden of disease** |  |  |  |  |
| Rare & mild | -0.198 (0.055; 0.266) | 70.146 | 3 | <0.0001 |
| Common & mild | -0.344 (0.042; 0.305) |  |  |  |
| Rare & severe | 0.187 (0.045; 0.178) |  |  |  |
| Common & severe | 0.355 (0.053; 0.216) |  |  |  |
| **Mild VRSE** |  |  |  |  |
| Common | -0.143 (0.026; 0.087) | 30.732 | 1 | <0.0001 |
| Rare | 0.143 (0.026; 0.085) |  |  |  |
| Note: Mean estimates corresponding to the last level of an attribute are calculated as minus the sum of the estimates for the other levels of the attribute. |  |  |  |  |

**Table 5.** Panel mixed logit model estimates (means and standard deviations) and significances of the attribute effects obtained from likelihood ratio (LR) tests. **France**

| Term | Mean estimate (std dev;<br>subject std dev) | LR Chi-Square | DF | P-value |
| --- | --- | --- | --- | --- |
| **‘Oneself’ model** |  |  |  |  |
| **Accessibility**<br>Co-payment & prescription<br>Free & accessible | -0.389 (0.026; 0.410)<br>0.389 (0.025; 0.383) | 238.254 | 1 | <0.0001 |
| **Vaccine effectiveness**<br>50%<br>90% | -0.375 (0.030; 0.222)<br>0.375 (0.032; 0.218) | 131.380 | 1 | <0.0001 |
| **Burden of disease**<br>Rare & mild<br>Common & mild<br>Rare & severe<br>Common & severe | -0.364 (0.052; 0.318)<br>-0.358 (0.042; 0.416)<br>0.273 (0.048; 0.224)<br>0.449 (0.053; 0.187) | 122.873 | 3 | <0.0001 |
| **Population coverage (x10%)** | 0.079 (0.010; 0.131) | 48.157 | 1 | <0.0001 |
| **Mild VRSE**<br>Common<br>Rare | -0.164 (0.027; 0.092)<br>0.164 (0.025; 0.093) | 44.450 | 1 | <0.0001 |
| **Local coverage (x10%)** | 0.064 (0.010; 0.093) | 33.480 | 1 | <0.0001 |
| **‘Youngest child’ model** |  |  |  |  |
| **Burden of disease**<br>Rare & mild<br>Common & mild<br>Rare & severe<br>Common & severe | -0.369 (0.048; 0.300)<br>-0.474 (0.051; 0.323)<br>0.331 (0.049; 0.202)<br>0.512 (0.048; 0.190) | 163.809 | 3 | <0.0001 |
| **Vaccine effectiveness**<br>50%<br>90% | -0.430 (0.029; 0.231)<br>0.430 (0.034; 0.237) | 152.182 | 1 | <0.0001 |
| **Accessibility**<br>Co-payment & prescription<br>Free & accessible | -0.314 (0.023; 0.278)<br>0.314 (0.025; 0.260) | 144.967 | 1 | <0.0001 |
| **Population coverage (x10%)** | 0.108 (0.010; 0.135) | 88.489 | 1 | <0.0001 |
| **Mild VRSE**<br>Common<br>Rare | -0.180 (0.026; 0.098)<br>0.180 (0.022; 0.095) | 46.913 | 1 | <0.0001 |
| **Local coverage (x10%)** | 0.078 (0.010; 0.086) | 44.981 | 1 | <0.0001 |
| Note: Mean estimates corresponding to the last level of an attribute are calculated as minus the sum of the estimates for the other levels of the attribute. |  |  |  |  |

Vaccine effectiveness stands out as the most important attribute in the UK, South Africa and in the ‘child’ group in the Netherlands. In Belgium and France, we found vaccine effectiveness to be a crucial element as well, at a relative importance of about 80% and 60% respectively. In all countries, vaccine effectiveness was ranked more important, or equally important, in the ‘child’ group compared to the ‘oneself’ group.

In contrast, vaccine accessibility was valued higher, or equally, in the ‘oneself’ group compared to the ‘child’ group in all countries, except for South Africa. In addition, accessibility was the most important attribute in Belgium, and in the ‘oneself’ group in France and the Netherlands.

The ‘child’ group in France cared most about the burden of the disease. This attribute was also of considerable importance in the same group in the Netherlands. There are notable differences in valuation of this attribute between all subgroups involved. Indeed, in the UK sample, the burden of disease was valued at a relative importance of about 40% in the ‘oneself’ group and about 20% in the ‘child’ group. Whereas in the Netherlands, France and Belgium, this attribute was valued at a relative importance of about 50% or more.

We observed a clear distinction between the ‘oneself’ group and the ‘child’ group with respect to population coverage. Indeed, both these indicators of vaccination coverage were considered relatively more important for children than for adults in all countries except the Netherlands. This implies that when parents decide about vaccinating their child, they are more prone to peer influence, than when adults (including parents) make these decisions for themselves. Overall, both population and local coverage were considered most important in France and South Africa and least important in the Netherlands and the UK.

Mild VRSE and local coverage were, although statistically significant for all subgroups, found to be of the relative lowest importance in most countries and subgroups.

Attribute-level utility estimates are listed in Tables 3 to 5. As could be expected, respondents in all study populations preferred the most a vaccine with 90% effectiveness, that is free & accessible, protects against a common & severe disease, rarely exhibits mild VRSE and for which vaccination coverage is high. In addition, we consistently found disease severity to dominate frequency of disease in all study samples.

The models including the attributes’ main effects as well as the most important covariate interactions are provided in Appendix A. Vaccine-related attitude statements were able to explain most preference heterogeneity. For example for Belgium, in the ‘oneself’ model we discovered respondents agreeing (disagreeing) with the statement “The available vaccinations are suited to protect my health” attached more (less) value to a vaccine with an efficacy of 90% (50%), compared to the average (see Figure S1 below). Moreover, we also found that in the ‘oneself’ model for the UK, respondents indicating that they were at low risk of contracting measles, cared more about the vaccine being free & accessible (see Figure S3 above). The same is true for individuals agreeing with “vaccinating my child is the logical thing to do” in the ‘child’ model for the UK (see Figure S4 above). For details on additional covariate interactions, we refer to Appendix A.

## Discussion

The need for behavioral data in relation to infectious disease epidemiology and prevention has been raised repeatedly over the past decade [16–18]. Our multi-country series of DCEs generated highly valuable data for parameterization and validation of epidemiological models. This is because data-driven hosts’ behavior derived from DCEs can be added to models mimicking the spread of infectious diseases. For example, dynamic behavior can be modelled through a utility function using prevalence utility estimates from the burden of disease attribute. As such, the utility of a vaccine increases when a disease becomes more prevalent. Similar dynamics can be modelled using the utility estimates of population or local coverage. Moreover, exogenous shocks, such as changing risk perceptions, can be introduced in such integrated models. Utility estimates on vaccine effectiveness, accessibility, disease severity and mild VRSE can facilitate data-driven introductions of exogenous shocks. Furthermore, the multi-country character of our study allows modelling vaccination behavior in five countries.

However, an integrated model combining data-driven vaccination behavior with infectious disease transmission dynamics, requires the specification of a dichotomous vaccine outcome (to be vaccinated or not) based on individual utilities derived from vaccine attributes. That is, a function that derives vaccine uptake from utility associated with vaccination. Future research will explore the specification of such vaccine uptake functions.

The positive estimates for both coverage attributes in all (sub)samples imply social norms or peer influence are more important than free-rider incentives. These findings confirm the positive coverage estimates reported in vaccination DCEs in Australia [19] and in the US [26]. Overall, it seems unlikely that respondents take externalities - such as herd immunity - into account when making vaccine decisions. As such, game theoretical models characterizing vaccine decisions as a strategic interaction between rational individuals, seem inappropriate to capture real-life vaccination dynamics. If individuals do include herd immunity effects in their decisions, it might very well be the case that they behave altruistically and opt for vaccination, contributing to the protection of vulnerable individuals. This was observed in several empirical studies, such as the study by Skea et al. [41] reporting on ‘avoiding harm to others’ incentives in the context of MMR vaccination in the UK. They found parents on a chat forum to be critical towards parents not vaccinating their healthy children, thereby putting vulnerable ones at risk. Altruistic motives were also described in the papers by Hakim et al. [42] and Shim et al. [43] in the context of influenza vaccination, and by Vietri et al. [44] about assessing the extent of altruism with respect to human papillomavirus (HPV) and influenza vaccination. Policy-makers and healthcare workers can influence vaccine hesitant individuals by communicating high coverage levels, i.e. describing that “accepting the vaccine is the mainstream thing to do”, in addition to other strategies (see Leask et al. [45] for a framework on “communicating with parents about vaccination”).

Vaccine accessibility has proven highly significant in our study, as well as in other studies [19, 22, 24–26], where it was, however, mostly described in terms of out-of-pocket or total costs. For example, Wong et al. [46] performed a DCE on mothers’ preferences for HPV vaccination in Hong Kong and found a significant impact of out-of-pocket cost on the decision to receive the vaccine. Poulos et al. [47] reported similar results with respect to traveler vaccines. This has also been confirmed by observational studies. For instance, in a retrospective cohort study, Lefevere et al. [48] found both personal information letters and removing out-of-pocket costs had a significant positive effect on HPV vaccination initiation in Belgium.

Given the importance of vaccine accessibility, policy-makers can increase vaccine coverage by making vaccines easily available at an affordable price. There is still significant room for improvement concerning adults (cfr. the ‘oneself group’) who are often confronted with an expensive, complicated and time consuming process of vaccination. Take for instance influenza vaccination in Belgium, where individuals typically visit a General Practitioner (GP) for a prescription, then buy the vaccine (often without reimbursement) at a pharmacy and lastly have to go back to the GP to be vaccinated. Not surprisingly, influenza vaccine coverage has usually been below 25% [49]. Adults cannot rely on the routine vaccination services available for children (e.g. well-baby clinics, child health clinics or school health centres). In this respect, (expansion of) workplace vaccination can play a vital role in facilitating vaccination for working-age adults. Policy-makers should consider incentivizing employers to offer certain vaccines to their employees at the workplace, e.g. influenza, and tetanus, diphtheria and pertussis (Tdap) vaccination, or hepatitis A for employees working in the food industry. Workplace vaccination may also prove useful in catch-up campaigns which would, for example, be required to maintain measles elimination targets [50–52]. Note that for the ‘child’ group, accessibility was found to be very important as well. Policy-makers should remain focused on making vaccines as accessible as possible for both groups.

In line with previous studies [22, 24, 46], vaccine effectiveness was observed to be of great importance in all models. Therefore it is essential that the public should remain fully aware of the positive impact vaccines are having on population health. According to a 2018 Gallup poll, the effectiveness of vaccines is perceived to be significantly more reliable than their safety. Of the five countries included in our study, France scored worst with about 20% of the population disagreeing that vaccines are effective, followed by The Netherlands and South Africa both at 11% [11].

In a previous study in Flanders [23], we applied the same DCE design but did not specify the severity of VRSE. The updated description in this study, varying only mild VRSE and describing severe VRSE as being ‘highly unlikely’, shifted the attribute’s importance from the highest rank in the earlier study, to one of the lowest ranks in the current. Safety concerns with respect to vaccinations remain crucial in vaccine misperceptions. VRSE may indeed occur, but are mostly mild and clear up quickly [53]. In this study it became clear that when respondents used realistic information on vaccine side-effects they cared less about them while making vaccination decisions. Acknowledging the existence of VRSE and providing risk and benefit information is recommended when discussing safety concerns with potential vaccine recipients (or their parents) [45]. The relative importance of burden of disease is more volatile and appears to be country-specific. In the ‘child’ model for the UK, we found it to be relatively unimportant, whereas for the same subpopulation in France, burden of disease was most important. The severity of the infectious disease was found to be more important than the frequency of the disease. This is in line with Sadique et al. [21], who showed severity of both vaccine-preventable disease and VRSE to be more important than their frequency. To address concerns about the burden of disease and VRSE, healthcare workers as key informants, should be well-versed in the general topic of vaccination and should use standard guidelines for each vaccine and disease so that potential vaccine recipients are consistently and well-informed. See also Leask et al. [45], who provide a vaccine communication framework.

## Conclusions

In conclusion, we found slightly different preferences for vaccine attributes between countries. Nonetheless, there are communalities in that people’s vaccine decisions seem to depend in the first place on how they perceive the effectiveness and risks of severe VRSE, as well as the burden of vaccine preventable disease. Their decisions are also influenced significantly by how easy it is to be vaccinated, in terms of effort and costs, by the possibility of mild VRSE and by how many other people are being vaccinated. Especially vaccination of the population in general is an important element when having a child vaccinated. Therefore communication strategies on vaccination should not forget to include information on vaccination rates, reflecting that vaccination is still the norm, and non-vaccination remains exceptional. Contrary to what most game theoretical models assume, this information would be an incentive to receive vaccination, rather than to forego it intending to take a free ride.

## Data Availability

The informed consent form specified that survey responses would only be made available and be analyzed by researchers at the University of Antwerp. As such, we cannot make the dataset with survey responses available to third parties.

## List of abbreviations

DCE: Discrete Choice Experiment
ECDC: European Centre for Disease Prevention and Control
EU: European Union
FDA: Food and Drug Administration
GP: General Practitioner
HPV: Human papillomavirus
LR: Likelihood Ratio
MMR: Measles – Mumps – Rubella
PML: Panel Mixed Logit
St dev: Standard deviation
UK: United Kingdom
VRSE: Vaccine related side-effects

## Declarations

### Ethics approval and consent to participate

The multi-country study protocol as well as the amendments to the country-specific questionnaires (Reference number: 15/2/12) were reviewed and accepted by the Ethical Committee of the Antwerp University Hospital (UZA, Belgium). The study was conducted according to the original approved protocol. Given that this study was non-interventional, and carried out in a general population (adults only) with informed consent, duplicate ethical approval at a local committee was not deemed necessary by our institutional review board. This was confirmed by an NHS Health Research Authority and Medical Research Council tool (UK) as well as by the Comité d’Ethique de Recherche en Maladies Infectieuses et Tropicales (France). Participants consented to the study protocol by continuing the questionnaire after the introduction stating the declaration of anonymity and voluntary participation. By continuing the questionnaire participants agreed with the processing and analysis of their data.

### Consent for publication

Not applicable.

### Authors’ contributions

PB initiated the study. FV, RK, LW and PB conceived and designed the questionnaire. FV and LW cleaned the raw data. FV and RK analyzed the data and constructed the models. All authors reviewed the manuscript.

## Acknowledgements

We thank all respondents who participated in the study. We also thank Wim Delva, Joram Hoogink, Albert Jan van Hoek, Aura Timen, Jacco Wallinga and Ardine de Wit for their substantial efforts in the South African and Dutch studies.

## Additional files

“Additional file 1.pdf” contains supporting information:

**Appendix A. PML model estimates including covariate interaction effects**

**Appendix B. Likert scale responses to vaccine attitude statements**

“Additional file 2.pdf” contains the questionnaire

## References

[1] World Health Organization. Health Topics — Immunization. Available from: https://www.who.int/topics/immunization/en/ (2019). Accessed 8 Aug 2019.

[2] Godlee F, Smith J, Marcovitch H. Wakefield’s article linking MMR vaccine and autism was fraudulent. British Medical Journal. 2011;c7452.

[3] Kata A. A postmodern Pandora’s box: anti-vaccination misinformation on the Internet. Vaccine. 2010;28(7):1709–1716.

[4] Ward JK, Peretti-Watel P, Larson HJ, Raude J, Verger P. Vaccine-criticism on the internet: new insights based on French-speaking websites. Vaccine. 2015;33(8):1063–1070.

[5] Larson HJ, de Figueiredo A, Xiahong Z, Schulz WS, Verger P, Johnston IG, et al. The state of vaccine confidence 2016: global insights through a 67-country survey. EBioMedicine. 2016;12:295–301.

[6] Hulsey E, Bland T. Immune overload: Parental attitudes toward combination and single antigen vaccines. Vaccine. 2015;33(22):2546–2550.

[7] Ruijs WL, Hautvast JL, van der Velden K, de Vos S, Knippenberg H, Hulscher ME. Religious subgroups influencing vaccination coverage in the Dutch Bible belt: an ecological study. BMC Public Health. 2011;11(1):102.

[8] Spier RE. Perception of risk of vaccine adverse events: a historical perspective. Vaccine. 2001;20:S78–S84.

[9] Poland GA, Jacobson RM. The age-old struggle against the antivaccinationists. New England Journal of Medicine. 2011;364(2):97–99.

[10] Larson HJ, Cooper LZ, Eskola J, Katz SL, Ratzan S. Addressing the vaccine confidence gap. The Lancet. 2011;378(9790):526–535.

[11] Gallup. Wellcome Global Monitor - First Wave Findings. Wellcome Trust; 2019.

[12] Centers for Disease Control and Prevention. Measles, Mumps, and Rubella (MMR) Vaccine Safety; 2018. Available from: https://www.cdc.gov/vaccinesafety/vaccines/mmr-vaccine.html (2018). Accessed 19 Dec 2019.

[13] European Centre for Disease Prevention and Control. Who is at risk for measles in the EU/EEA? Identifying susceptible groups to close immunity gaps towards measles elimination. ECDC; 2019.

[14] Nelson R. US measles outbreak concentrated among unvaccinated children. The Lancet Infectious Diseases. 2019;19(3):248.

[15] Patel M. Increase in measles cases? United States, January 1–April 26, 2019. MMWR Morbidity and Mortality Weekly Report. 2019;68.

[16] Verelst F, Willem L, Beutels P. Behavioural change models for infectious disease transmission: a systematic review (2010–2015). Journal of The Royal Society Interface. 2016;13(125):20160820.

[17] Funk S, Salathé M, Jansen VA. Modelling the influence of human behaviour on the spread of infectious diseases: a review. Journal of the Royal Society Interface. 2010;7(50):1247–1256.

[18] Funk S, Bansal S, Bauch CT, Eames KT, Edmunds WJ, Galvani AP, et al. Nine challenges in incorporating the dynamics of behaviour in infectious diseases models. Epidemics. 2015;10:21–25.

[19] Hall J, Kenny P, King M, Louviere J, Viney R, Yeoh A. Using stated preference discrete choice modelling to evaluate the introduction of varicella vaccination. Health Economics. 2002;11(5):457–465.

[20] de Bekker-Grob Ew, Hofman R, Donkers B, van Ballegooijen M, Helmerhorst TJ, Raat H, et al. Girls’ preferences for HPV vaccination: a discrete choice experiment. Vaccine. 2010;28(41):6692–6697.

[21] Sadique MZ, Devlin N, Edmunds WJ, Parkin D. The effect of perceived risks on the demand for vaccination: results from a discrete choice experiment. PLoS One. 2013;8(2):e54149.

[22] Determann D, Korfage IJ, Lambooij MS, Bliemer M, Richardus JH, Steyerberg EW, et al. Acceptance of vaccinations in pandemic outbreaks: a discrete choice experiment. PloS one. 2014;9(7):e102505.

[23] Verelst F, Willem L, Kessels R, Beutels P. Individual decisions to vaccinate one’s child or oneself: A discrete choice experiment rejecting free-riding motives. Social Science & Medicine. 2018;207:106–116.

[24] Verelst F, Kessels R, Delva W, Beutels P, Willem L. Drivers of vaccine decision-making in South Africa: A discrete choice experiment. Vaccine. 2019;37(15):2079–2089.

[25] Bishai D, Brice R, Girod I, Saleh A, Ehreth J. Conjoint analysis of French and German parents’ willingness to pay for meningococcal vaccine. Pharmacoeconomics. 2007;25(2):143–154.

[26] Gidengil C, Lieu TA, Payne K, Rusinak D, Messonnier M, Prosser LA. Parental and societal values for the risks and benefits of childhood combination vaccines. Vaccine. 2012;30(23):3445–3452.

[27] de Bekker-Grob EW, Ryan M, Gerard K. Discrete choice experiments in health economics: a review of the literature. Health Economics. 2012;21(2):145–172.

[28] Kessels R, Jones B, Goos P, Vandebroek M. The usefulness of Bayesian optimal designs for discrete choice experiments. Applied Stochastic Models in Business and Industry. 2011;27(3):173–188.

[29] Oteng B, Marra F, Lynd LD, Ogilvie G, Patrick D, Marra CA. Evaluating societal preferences for human papillomavirus vaccine and cervical smear test screening programme. Sexually Transmitted Infections. 2011;87(1):52–57.

[30] de Bekker-Grob EW, Swait JD, Kassahun HT, Bliemer MC, Jonker MF, Veldwijk J, et al. Are healthcare choices predictable? The impact of discrete choice experiment designs and models. Value in Health. 2019;22(9):1050–1062.

[31] Hoogink J, Verelst F, Kessels R, van Hoek AJ, Timen A, Willem L, et al. Preferential differences in vaccination decision-making for oneself or one’s child in The Netherlands: a discrete choice experiment. BMC Public Health. 2020;20:828.

[32] Peretti-Watel P, Verger P, Raude J, Constant A, Gautier A, Jestin C, et al. Dramatic change in public attitudes towards vaccination during the 2009 influenza A (H1N1) pandemic in France. Eurosurveillance. 2013;18(44):20623.

[33] Amirthalingam G, Gupta S, Campbell H. Pertussis immunisation and control in England and Wales, 1957 to 2012: a historical review. Eurosurveillance. 2013;18(38):20587.

[34] Vandermeulen C, Hoppenbrouwers K, Roelants M, Theeten H, Braeckman T, Maertens K, et al. Studie van de vaccinatiegraad in Vlaanderen 2016; 2017.

[35] Tjalma W, Brasseur C, Top G, Ribesse N, Morales I, Van Damme P. HPV vaccination coverage in the federal state of Belgium according to regions and their impact. Facts, views & vision in ObGyn. 2018;10(2):101.

[36] Bults M, Beaujean DJ, de Zwart O, Kok G, van Empelen P, van Steenbergen JE, et al. Perceived risk, anxiety, and behavioural responses of the general public during the early phase of the InfluenzaA (H1N1) pandemic in the Netherlands: results of three consecutive online surveys. BMC Public Health. 2011;11(1):2.

[37] Chew LD, Bradley KA, Boyko EJ. Brief questions to identify patients with inadequate health literacy. Family Medicine. 2004;36(8):588–94.

[38] SAS Institute Inc, Cary, NC. JMP;.

[39] RStudio Team. RStudio: Integrated Development Environment for R. Boston, MA; 2015. Available from: http://www.rstudio.com/.

[40] European Statistical System (ESS). 2011 Census Hub; 2011. Available from: https://ec.europa.eu/eurostat/web/population-and-housing-census/census-data/2011-census.

[41] Skea ZC, Entwistle VA, Watt I, Russell E. ‘Avoiding harm to others’ considerations in relation to parental measles, mumps and rubella (MMR) vaccination discussions–An analysis of an online chat forum. Social Science & Medicine. 2008;67(9):1382–1390.

[42] Hakim H, Gaur AH, McCullers JA. Motivating factors for high rates of influenza vaccination among healthcare workers. Vaccine. 2011;29(35):5963–5969.

[43] Shim E, Chapman GB, Townsend JP, Galvani AP. The influence of altruism on influenza vaccination decisions. Journal of The Royal Society Interface. 2012;p. rsif20120115.

[44] Vietri JT, Li M, Galvani AP, Chapman GB. Vaccinating to help ourselves and others. Medical Decision Making. 2012;32(3):447–458.

[45] Leask J, Kinnersley P, Jackson C, Cheater F, Bedford H, Rowles G. Communicating with parents about vaccination: a framework for health professionals. BMC pediatrics. 2012;12(1):154.

[46] Wong CK, Man KK, Ip P, Kwan M, McGhee SM. Mothers’ preferences and willingness to pay for human papillomavirus vaccination for their daughters: A discrete choice experiment in Hong Kong. Value in Health. 2018;21(5):622–629.

[47] Poulos C, Curran D, Anastassopoulou A, De Moerlooze L. German travelers’ preferences for travel vaccines assessed by a discrete choice experiment. Vaccine. 2018;36(7):969–978.

[48] Lefevere E, Hens N, De Smet F, Beutels P. The impact of non-financial and financial encouragements on participation in non school-based human papillomavirus vaccination: a retrospective cohort study. The European Journal of Health Economics. 2016;17(3):305–315.

[49] Tafforeau J. Vaccinatie. In: Demarest S, Charafeddine R, editors. Gezondheidsenquête 2013. Rapport 5: Preventie. Brussel: WIV-ISP; 2015.

[50] Kuylen E, Willem L, Hens N, Broeckhove J. Future Ramifications of Age-Dependent Immunity Levels for Measles: Explorations in an Individual-Based Model. In: International Conference on Computational Science. Springer; 2019. p. 456–467.

[51] Funk S, Camacho A, Kucharski AJ, Eggo RM, Edmunds WJ. Real-time forecasting of infectious disease dynamics with a stochastic semi-mechanistic model. Epidemics. 2018;22:56–61.

[52] Trentini F, Poletti P, Merler S, Melegaro A. Measles immunity gaps and the progress towards elimination: a multi-country modelling analysis. The Lancet Infectious Diseases. 2017;17(10):1089–1097.

[53] World Health Organization. Global Vaccine Safety. Adverse events following immunization (AEFI). https://www.who.int/vaccine_safety/initiative/detection/AEFI/en/ (2019). Accessed 3 Aug 2020.

