## Additional file 1 for "No such thing as a free-rider? Understanding multicountry drivers of childhood and adult vaccination"

### Supporting Information for: "No such thing as a free-rider? Understanding multicountry drivers of childhood and adult vaccination"

Frederik Verelst[1*], Roselinde Kessels[2,3], Lander Willem[1], Philippe Beutels[1,4]

**1** Centre for Health Economics Research and Modelling Infectious Diseases, Vaccine and Infectious Disease Institute, University of Antwerp, Antwerp, Belgium

**2** Department of Economics & Flemish Research Foundation (FWO), University of Antwerp, Antwerp, Belgium

**3** Department of Data Analytics and Digitalization, Maastricht University, Maastricht, The Netherlands

**4** School of Public Health and Community Medicine, The University of New South Wales, Sydney, Australia

*Corresponding author: Frederik Verelst, Centre for Health Economics and Modelling Infectious Diseases (CHERMID), Vaccine & Infectious Disease Institute (VAXINFECTIO), University of Antwerp, Campus Drie Eiken, Room S2.42, Universiteitsplein 1, 2610 Wilrijk (Antwerpen), Belgium,

#### Contents

**Appendix A: PML model estimates including covariate interaction effects** **2**

**Appendix B: Likert scale responses to vaccine attitude statements** **14**

### Appendix A: PML model estimates including covariate interaction effects

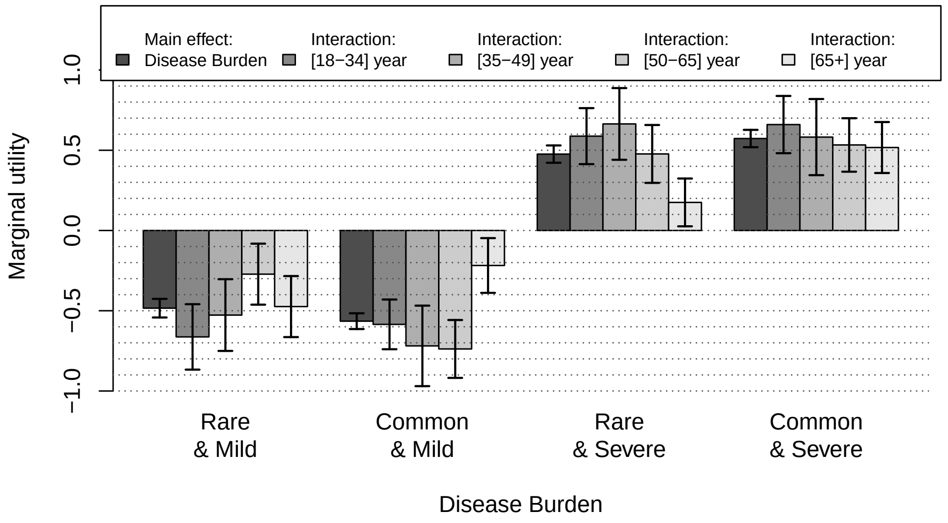


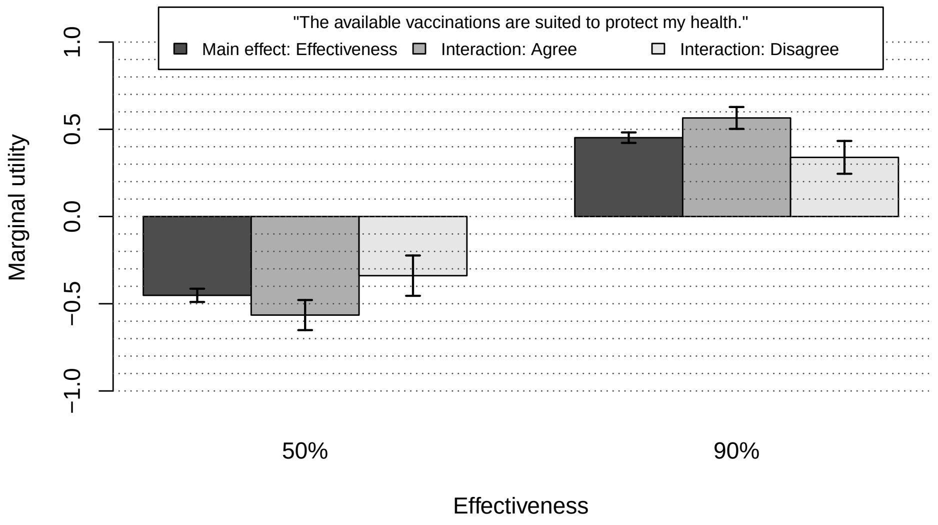


**Fig S1. Marginal utilities for significant covariate interactions with disease burden (above) and vaccine effectiveness (below). 'Oneself' model, Belgium.**

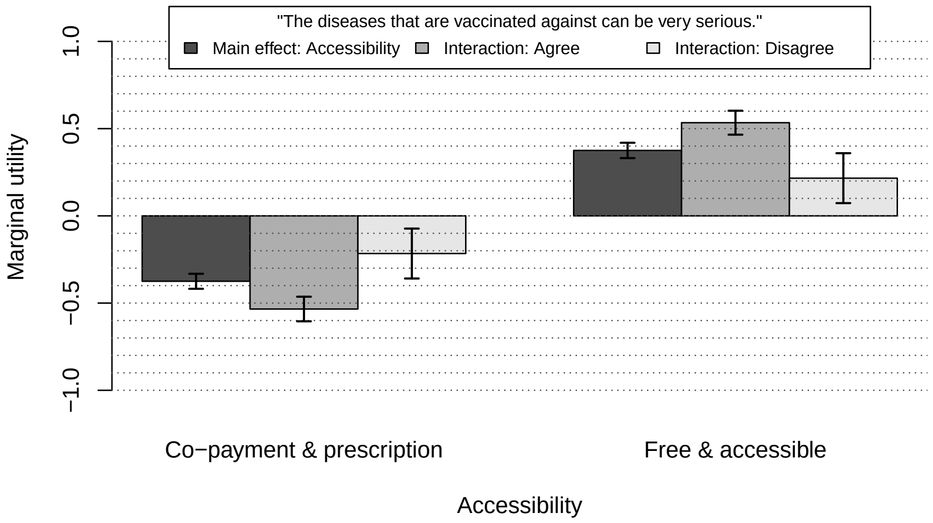


**Fig S2. Marginal utilities for significant covariate interaction with accessibility. 'Youngest child' model, Belgium.**

**Table S1. Panel mixed logit model estimates (means and standard deviations) and significances of the attribute effects obtained from likelihood ratio (LR) tests. 'Oneself' model Belgium, including covariate interactions**

| **Term** | **Mean estimate (std dev; subject std dev)** | **LR Chi-square** | **DF** | **P-value** |
|---|---|---|---|---|
| **Accessibility** | | | | |
| Co-payment & prescription | -0.433 (0.024; 0.376) | 327.736 | 1 | < 0.0001 |
| Free & accessible | 0.433 (0.022; 0.370) | | | |
| **Burden of disease** | | | | |
| Rare & mild | -0.484 (0.058; 0.187) | 257.965 | 3 | < 0.0001 |
| Common & mild | -0.565 (0.049; 0.132) | | | |
| Rare & severe | 0.476 (0.054; 0.099) | | | |
| Common & severe | 0.573 (0.054; 0.159) | | | |
| **Vaccine effectiveness** | | | | |
| 50% | -0.452 (0.038; 0.153) | 192.235 | 1 | < 0.0001 |
| 90% | 0.452 (0.030; 0.144) | | | |
| **Population coverage (x10%)** | 0.086 (0.009; 0.125) | 66.439 | 1 | < 0.0001 |
| **Mild VRSE** | | | | |
| Common | -0.179 (0.024; 0.112) | 58.630 | 1 | < 0.0001 |
| Rare | 0.179 (0.022; 0.101) | | | |
| **Vaccine effectiveness*Protective**[†] | | | | |
| 50%*agree | -0.113 (0.027; 0.162) | 18.087 | 1 | < 0.0001 |
| 50%*disagree | 0.113 (0.028; 0.162) | | | |
| 90%*agree | 0.113 (0.025; 0.127) | | | |
| 90%*disagree | -0.113 (0.024; 0.140) | | | |
| **Local coverage (x10%)** | 0.049 (0.009; 0.084) | 18.079 | 1 | < 0.0001 |
| **Burden of disease*Age** | | | | |
| Rare & mild*[18-34] | -0.179 (0.087; 0.210) | 33.439 | 9 | 0.0001 |
| Rare & mild*[35-49] | -0.043 (0.096; 0.251) | | | |
| Rare & mild*[50-65] | 0.212 (0.087; 0.254) | | | |
| Rare & mild*[65+] | 0.010 (0.105; 0.269) | | | |
| Common & mild*[18-34] | -0.020 (0.081; 0.174) | | | |
| Common & mild*[35-49] | -0.154 (0.099; 0.223) | | | |
| Common & mild*[50-65] | -0.173 (0.087; 0.200) | | | |
| Common & mild*[65+] | 0.347 (0.089; 0.246) | | | |
| Rare & severe*[18-34] | 0.112 (0.085; 0.140) | | | |
| Rare & severe*[35-49] | 0.188 (0.088; 0.146) | | | |
| Rare & severe*[50-65] | 0.001 (0.079; 0.170) | | | |
| Rare & severe*[65+] | -0.301 (0.068; 0.135) | | | |
| Common & severe*[18-34] | 0.087 (0.080; 0.146) | | | |
| Common & severe*[35-49] | 0.009 (0.095; 0.174) | | | |
| Common & severe*[50-65] | -0.040 (0.079; 0.185) | | | |
| Common & severe*[65+] | -0.056 (0.076; 0.143) | | | |

Note: Mean estimates corresponding to the last level of an attribute are calculated as minus the sum of the estimates for the other levels of the attribute. [†]Protective: "The available vaccinations are suited to protect my health."

**Table S2. Panel mixed logit model estimates (means and standard deviations) and significances of the attribute effects obtained from likelihood ratio (LR) tests. 'Youngest child' model Belgium, including covariate interactions**

| Term | Mean estimate (std dev; subject std dev) | LR Chi-square | DF | P-value |
|---|---|---|---|---|
| **Vaccine effectiveness** | | | | |
| 50% | -0.584 (0.033; 0.238) | 192.237 | 1 | < 0.0001 |
| 90% | 0.584 (0.041; 0.230) | | | |
| **Burden of disease** | | | | |
| Rare & mild | -0.380 (0.063; 0.342) | 163.220 | 3 | < 0.0001 |
| Common & mild | -0.621 (0.064; 0.458) | | | |
| Rare & severe | 0.359 (0.054; 0.256) | | | |
| Common & severe | 0.642 (0.068; 0.351) | | | |
| **Accessibility** | | | | |
| Co-payment & prescription | -0.375 (0.043; 0.182) | 94.231 | 1 | < 0.0001 |
| Free & accessible | 0.375 (0.044; 0.177) | | | |
| **Population coverage (x10%)** | 0.132 (0012.; 0.138) | 93.690 | 1 | < 0.0001 |
| **Mild VRSE** | | | | |
| Common | -0.229 (0.030; 0.140) | 45.361 | 1 | < 0.0001 |
| Rare | 0.229 (0.029; 0.134) | | | |
| **Local coverage (x10%)** | 0.074 (0.014; 0.120) | 27.390 | 1 | < 0.0001 |
| **Accessibility*Severity VPD†** | | | | |
| Co-payment & prescription*agree | -0.159 (0.042; 0.178) | 18.556 | 1 | < 0.0001 |
| Co-payment & prescription*disagree | 0.159 (0.037; 0.179) | | | |
| Free & accessible*agree | 0.159 (0.037; 0.185) | | | |
| Free & accessible*disagree | -0.159 (0.037; 0.175) | | | |

Note: Mean estimates corresponding to the last level of an attribute are calculated as minus the sum of the estimates for the other levels of the attribute. †Severity VPD: "The diseases that are vaccinated against can be very serious."

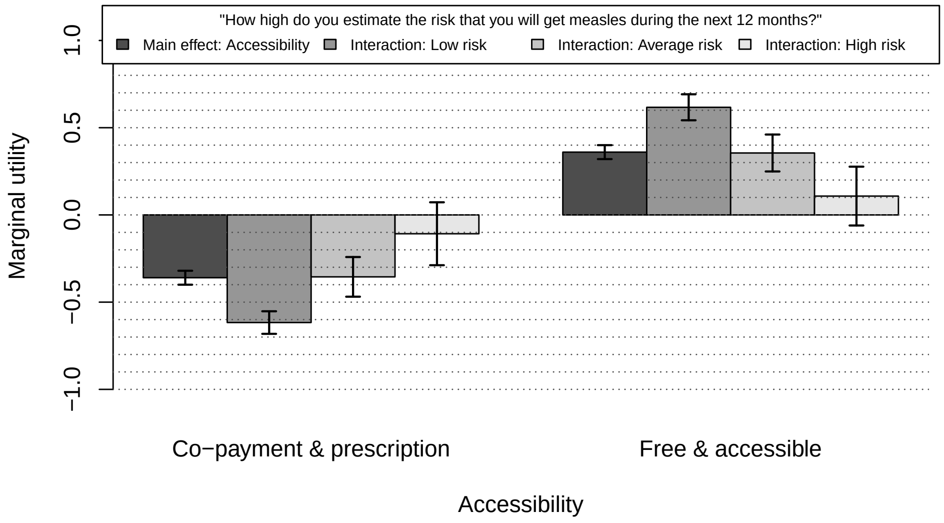


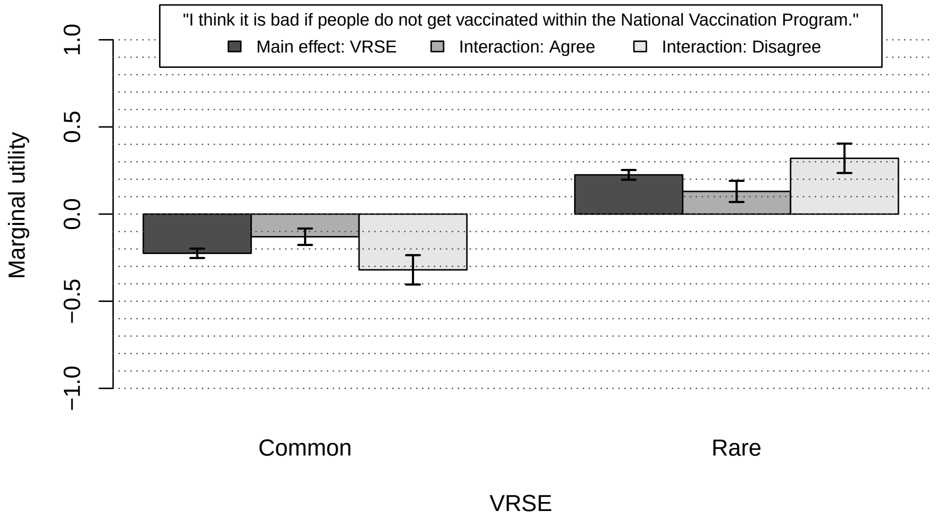


**Fig S3. Marginal utilities for significant covariate interactions with accessibility (above) and VRSE (below). 'Oneself' model, United Kingdom.**

**Table S3. Panel mixed logit model estimates (means and standard deviations) and significances of the attribute effects obtained from likelihood ratio (LR) tests. 'Oneself' model United Kingdom, including covariate interactions**

| Term | Mean estimate (std dev; subject std dev) | LR Chi-square | DF | P-value |
|---|---|---|---|---|
| **Vaccine effectiveness** | | | | |
| 50%<br>90% | -0.720 (0.037; 0.301)<br>0.720 (0.034; 0.296) | 431.561 | 1 | < 0.0001 |
| **Burden of disease** | | | | |
| Rare & mild<br>Common & mild<br>Rare & severe<br>Common & severe | -0.545 (0.053; 0.310)<br>-0.368 (0.055; 0.438)<br>0.327 (0.046; 0.219)<br>0.586 (0.056; 0.259) | 192.478 | 3 | < 0.0001 |
| **Accessibility** | | | | |
| Co-payment & prescription<br>Free & accessible | -0.360 (0.040; 0.124)<br>0.360 (0.040; 0.120) | 121.213 | 1 | < 0.0001 |
| **Population coverage (x10%)** | 0.099 (0.010; 0.128) | 94.070 | 1 | < 0.0001 |
| **Mild VRSE** | | | | |
| Common<br>Rare | -0.225 (0.027; 0.087)<br>0.225 (0.028; 0.089) | 66.328 | 1 | < 0.0001 |
| **Local coverage (x10%)** | 0.084 (0.010; 0.080) | 48.389 | 1 | < 0.0001 |
| **Accessibility*Measles susceptibility†** | | | | |
| Co-payment & prescription*low risk<br>Co-payment & prescription*average risk<br>Co-payment & prescription*high risk<br>Free & accessible*low risk<br>Free & accessible*average risk<br>Free & accessible*high risk | -0.257 (0.042; 0.167)<br>0.005 (0.050; 0.143)<br>0.252 (0.067; 0.133)<br>0.257 (0.045; 0.168)<br>-0.005 (0.046; 0.129)<br>-0.252 (0.061; 0.134) | 39.611 | 2 | < 0.0001 |
| **Mild VRSE*Bad if others don't vaccinate‡** | | | | |
| Common*agree<br>Common*disagree<br>Rare*agree<br>Rare*disagree | 0.095 (0.022; 0.091)<br>-0.095 (0.026; 0.093)<br>-0.095 (0.025; 0.088)<br>0.095 (0.025; 0.094) | 17.408 | 1 | < 0.0001 |

Note: Mean estimates corresponding to the last level of an attribute are calculated as minus the sum of the estimates for the other levels of the attribute. †Measles susceptibility: "How high do you estimate the risk that you will get measles during the next 12 months?" ‡Bad if others don't vaccinate: "I think it is bad if people do not get vaccinated within the National Vaccination Program."

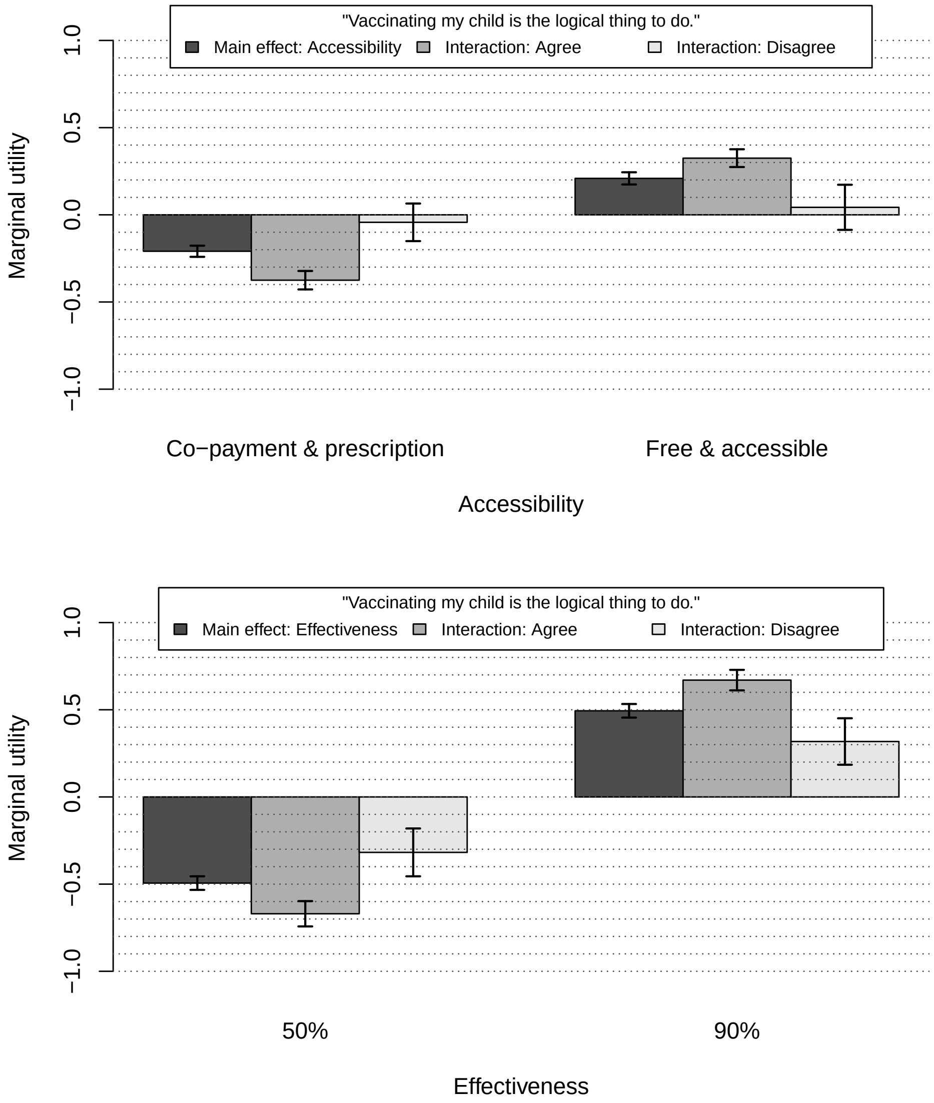


**Fig S4. Marginal utilities for significant covariate interactions with accessibility (above) and vaccine effectiveness (below). 'Youngest child' model, United Kingdom.**

**Table S4. Panel mixed logit model estimates (means and standard deviations) and significances of the attribute effects obtained from likelihood ratio (LR) tests. 'Youngest child' model United Kingdom, including covariate interactions**

| Term | Mean estimate (std dev; subject std dev) | LR Chi-square | DF | P-value |
|---|---|---|---|---|
| **Vaccine effectiveness** | | | | |
| 50%<br>90% | -0.494 (0.039; 0.159)<br>0.494 (0.039; 0.152) | 144.973 | 1 | < 0.0001 |
| **Population coverage (x10%)** | 0.112 (0.010; 0.107) | 95.334 | 1 | < 0.0001 |
| **Local coverage (x10%)** | 0.100 (0.012; 0.100) | 68.070 | 1 | < 0.0001 |
| **Burden of disease** | | | | |
| Rare & mild<br>Common & mild<br>Rare & severe<br>Common & severe | -0.204 (0.048; 0.275)<br>-0.359 (0.046; 0.316)<br>0.198 (0.050; 0.174)<br>0.365 (0.046; 0.223) | 71.456 | 3 | < 0.0001 |
| **Accessibility** | | | | |
| Co-payment & prescription<br>Free & accessible | -0.209 (0.032; 0.140)<br>0.209 (0.035; 0.130) | 40.391 | 1 | < 0.0001 |
| **Mild VRSE** | | | | |
| Common<br>Rare | -0.146 (0.025; 0.094)<br>0.146 (0.023; 0.092) | 31.056 | 1 | < 0.0001 |
| **Vaccine effectiveness*Logical†** | | | | |
| 50%*agree<br>50%*disagree<br>90%*agree<br>90%*disagree | -0.176 (0.036; 0.137)<br>0.176 (0.038; 0.130)<br>0.176 (0.036; 0.142)<br>-0.176 (0.036; 0.136) | 26.399 | 1 | < 0.0001 |
| **Accessibility*Logical†** | | | | |
| Co-payment & prescription*agree<br>Co-payment & prescription*disagree<br>Free & accessible*agree<br>Free & accessible*disagree | -0.166 (0.035; 0.132)<br>0.166 (0.030; 0.134)<br>0.166 (0.036; 0.136)<br>-0.166 (0.035; 0.127) | 25.668 | 1 | < 0.0001 |

Note: Mean estimates corresponding to the last level of an attribute are calculated as minus the sum of the estimates for the other levels of the attribute. †Logical: "Vaccinating my child is the logical thing to do."

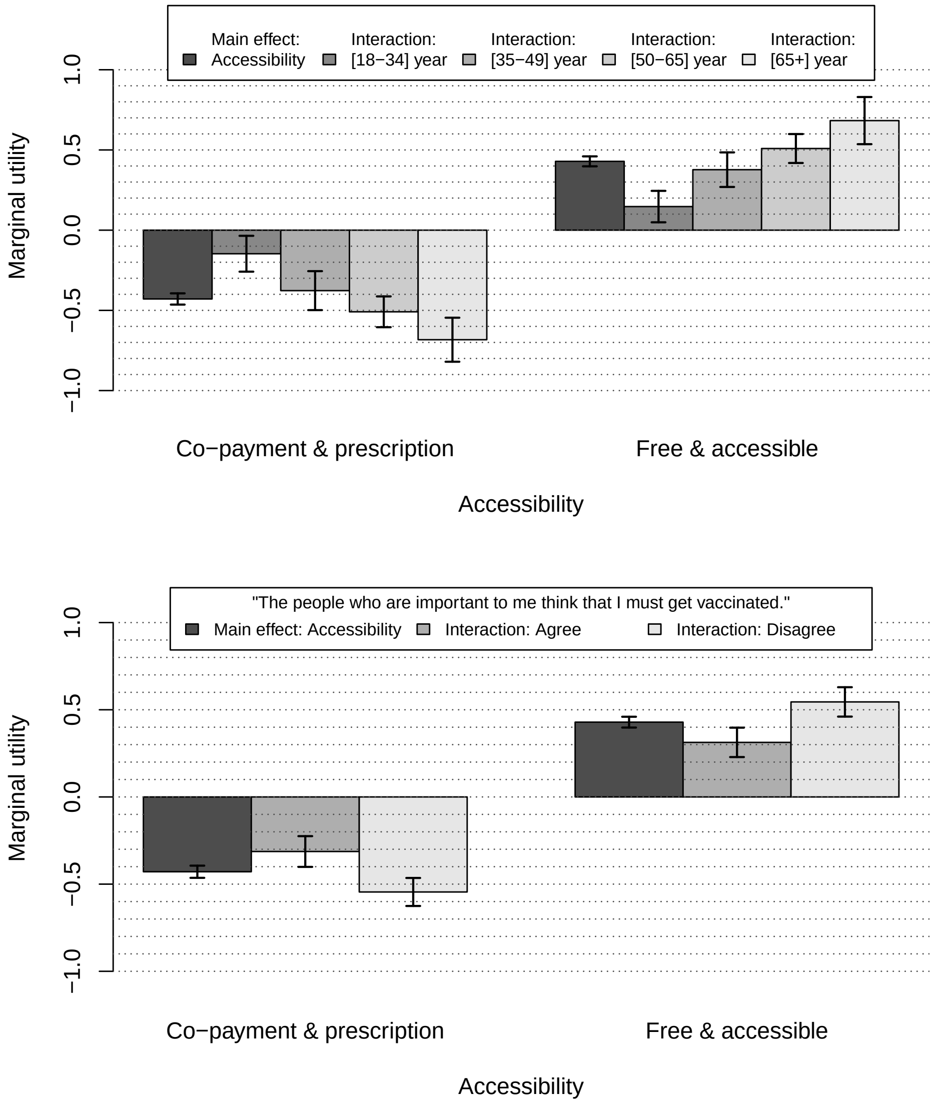


**Fig S5. Marginal utilities for significant covariate interactions with accessibility (both charts). 'Oneself' model, France.**

**Table S5. Panel mixed logit model estimates (means and standard deviations) and significances of the attribute effects obtained from likelihood ratio (LR) tests. 'Oneself' model France, including covariate interactions**

| Term | Mean estimate (std dev; subject std dev) | LR Chi-square | DF | P-value |
|---|---|---|---|---|
| **Accessibility** | | | | |
| Co-payment & prescription | -0.429 (0.035; 0.115) | 215.395 | 1 | < 0.0001 |
| Free & accessible | 0.429 (0.031; 0.106) | | | |
| **Vaccine effectiveness** | | | | |
| 50% | -0.403 (0.030; 0.237) | 137.847 | 1 | < 0.0001 |
| 90% | 0.403 (0.031; 0.228) | | | |
| **Burden of disease** | | | | |
| Rare & mild | -0.409 (0.051; 0.399) | 125.353 | 3 | < 0.0001 |
| Common & mild | -0.358 (0.048; 0.455) | | | |
| Rare & severe | 0.295 (0.050; 0.260) | | | |
| Common & severe | 0.472 (0.052; 0.231) | | | |
| **Mild VRSE** | | | | |
| Common | -0.184 (0.025; 0.097) | 48.017 | 1 | < 0.0001 |
| Rare | 0.184 (0.025; 0.102) | | | |
| **Population coverage (x10%)** | 0.083 (0.011; 0.143) | 47.914 | 1 | < 0.0001 |
| **Accessibility*Age** | | | | |
| Co-payment & prescription*[18-34] | 0.282 (0.060; 0.092) | 42.015 | 3 | < 0.0001 |
| Co-payment & prescription*[35-49] | 0.052 (0.052; 0.113) | | | |
| Co-payment & prescription*[50-65] | -0.080 (0.045; 0.159) | | | |
| Co-payment & prescription*[65+] | -0.254 (0.052; 0.157) | | | |
| Free & accessible*[18-34] | -0.282 (0.050; 0.092) | | | |
| Free & accessible*[35-49] | -0.052 (0.049; 0.120) | | | |
| Free & accessible*[50-65] | 0.080 (0.050; 0.174) | | | |
| Free & accessible*[65+] | 0.254 (0.058; 0.149) | | | |
| **Local coverage (x10%)** | 0.069 (0.010; 0.098) | 34.527 | 1 | < 0.0001 |
| **Accessibility*Peer influence†** | | | | |
| Co-payment & prescription*agree | 0.116 (0.026; 0.118) | 27.529 | 1 | < 0.0001 |
| Co-payment & prescription*disagree | -0.116 (0.028; 0.109) | | | |
| Free & accessible*agree | -0.116 (0.028; 0.110) | | | |
| Free & accessible*disagree | 0.116 (0.027; 0.107) | | | |

Note: Mean estimates corresponding to the last level of an attribute are calculated as minus the sum of the estimates for the other levels of the attribute. †Peer influence: "The people who are important to me think that I must get vaccinated."

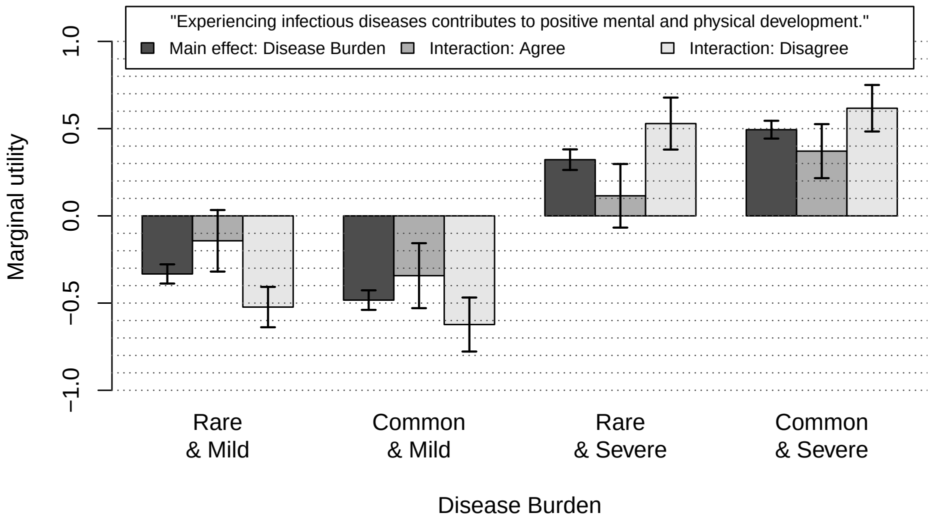


**Fig S6. Marginal utilities for significant covariate interaction with burden of disease. 'Youngest child' model, France.**

**Table S6. Panel mixed logit model estimates (means and standard deviations) and significances of the attribute effects obtained from likelihood ratio (LR) tests. 'Youngest child' model France, including covariate interactions**

| Term | Mean estimate (std dev; subject std dev) | LR Chi-square | DF | P-value |
|---|---|---|---|---|
| **Vaccine effectiveness** | | | | |
| 50% | -0.450 (0.028; 0.233) | 151.953 | 1 | < 0.0001 |
| 90% | 0.450 (0.031; 0.273) | | | |
| **Accessibility** | | | | |
| Co-payment & prescription | -0.330 (0.024; 0.316) | 145.020 | 1 | < 0.0001 |
| Free & accessible | 0.333 (0.023; 0.296) | | | |
| **Burden of disease** | | | | |
| Rare & mild | -0.333 (0.055; 0.249) | 124.816 | 3 | < 0.0001 |
| Common & mild | -0.483 (0.056; 0.227) | | | |
| Rare & severe | 0.322 (0.059; 0.178) | | | |
| Common & severe | 0.494 (0.051; 0.177) | | | |
| **Population coverage (x10%)** | 0.092 (0.014; 0.090) | 47.444 | 1 | < 0.0001 |
| **Mild VRSE** | | | | |
| Common | -0.187 (0.024; 0.112) | 47.161 | 1 | < 0.0001 |
| Rare | 0.187 (0.024; 0.104) | | | |
| **Local coverage (x10%)** | 0.081 (0.011; 0.092) | 45.150 | 1 | < 0.0001 |
| **Burden of disease*Positive development after infection†** | | | | |
| Rare & mild*agree | 0.190 (0.060; 0.216) | 27.754 | 3 | < 0.0001 |
| Rare & mild*disagree | -0.190 (0.055; 0.167) | | | |
| Common & mild*agree | 0.140 (0.058; 0.201) | | | |
| Common & mild*disagree | -0.140 (0.057; 0.200) | | | |
| Rare & severe*agree | -0.207 (0.055; 0.147) | | | |
| Rare & severe*disagree | 0.207 (0.055; 0.164) | | | |
| Common & severe*agree | -0.123 (0.047; 0.158) | | | |
| Common & severe*disagree | 0.123 (0.056; 0.376) | | | |
| **Population coverage (x10%)*Confidence in vaccine info‡** | | | | |
| Population coverage (x10%)*agree | 0.062 (0.014; 0.084) | 19.992 | 1 | < 0.0001 |
| Population coverage (x10%)*disagree | -0.062 (0.013; 0.087) | | | |

Note: Mean estimates corresponding to the last level of an attribute are calculated as minus the sum of the estimates for the other levels of the attribute. †Positive development after infection: "Experiencing infectious diseases contributes to positive mental and physical development." ‡Confidence in vaccine info: "I have confidence in the information about vaccinations that my care provider (*care provider is your GP or child healthcare professional/paediatrician) gives me."

### Appendix B: Likert scale responses to vaccine attitude statements

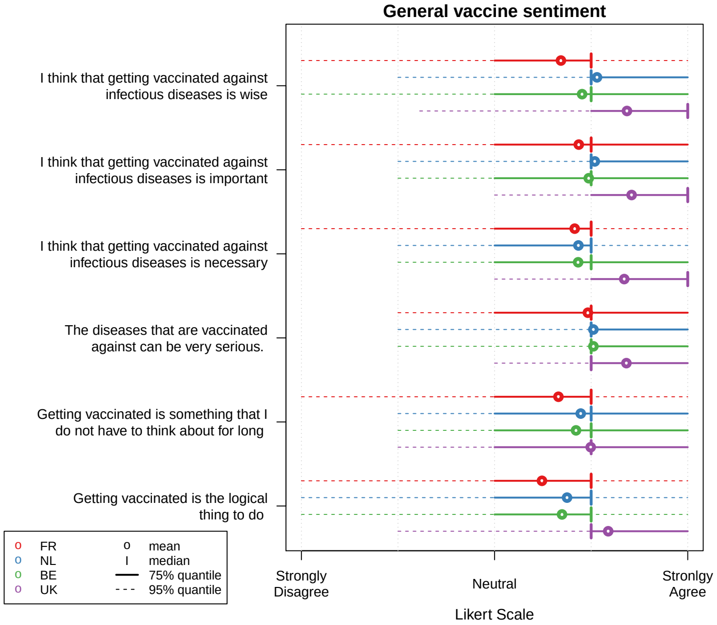


**Fig S7. Likert scale responses to general vaccine statements in the 'oneself' group**

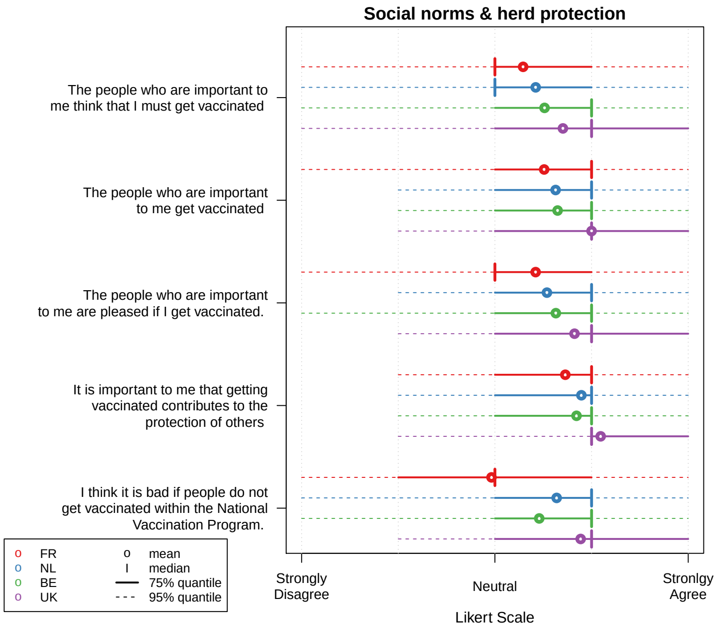


**Fig S8. Likert scale responses to statements with respect to social norms and herd protection in the 'oneself' group**

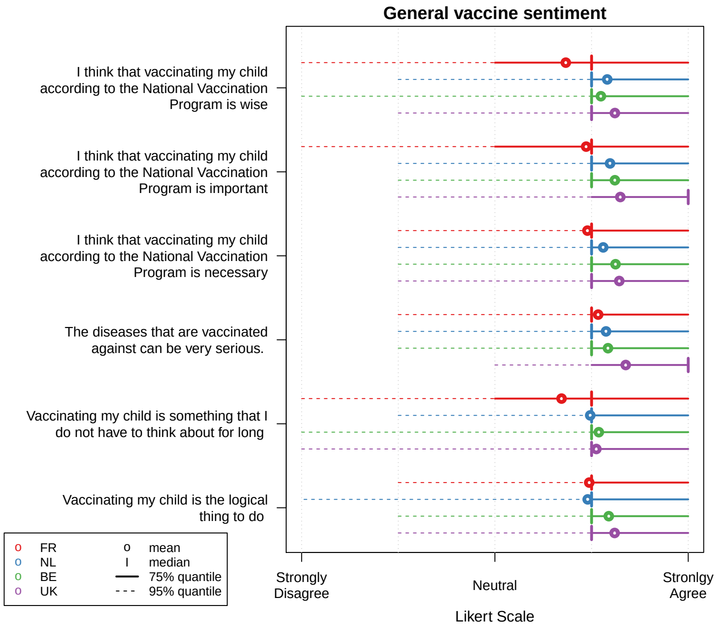


**Fig S9. Likert scale responses to general vaccine statements in the 'youngest child' group**

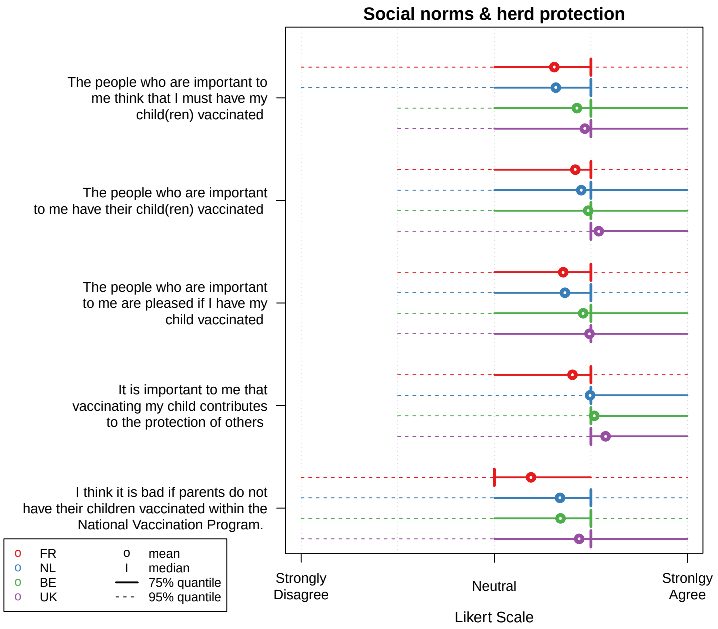


**Fig S10. Likert scale responses to statements with respect to social norms and herd protection in the ‘youngest child’ group**
