## Additional file 2 for "No such thing as a free-rider? Understanding multicountry drivers of childhood and adult vaccination"

**INFORMATION SHEET**

Dear Participant,

Please first read this information carefully before completing the questionnaire.

**What is the aim of this research?**

This questionnaire is part of a broader research programme conducted by the University of Antwerp and funded by the Flemish Fund for Scientific Research. With this research we want to find out what different people in Europe think about vaccination and which aspects about vaccinations are important for themselves, or, if they are a parent, for their children to decide whether or not to accept one of the vaccines in the National Immunization Programme.

The term vaccination is used in this questionnaire; this is also referred to as inoculations or jabs (such as in the flu jab). This survey is strictly about preventive vaccinations. Preventive vaccination is a way of **preventing** an infectious disease and is therefore **not** a medicine to treat a disease.

The first part of the questionnaire consists of some general questions about your background. In addition, we also ask you to respond to a number of statements, using a scale of 1 to 5 to indicate your opinion. The second part contains a total of 10 multiple choice questions in which you are presented with two vaccines that have different characteristics. Each time, we ask you to choose the vaccine that appeals to you the most. This questionnaire will take about 5-10 minutes in total.

**Participation and anonymity**

We emphasize that participation in this study is completely **voluntary and anonymous**. You can therefore stop your participation in this survey at any time, without any further consequences.

If you fill in and submit the survey, the data will be stored in a database by an independent marketing bureau. This database will be stripped of all potential identifiers and then sent to researchers at the University of Antwerp. The researchers will analyse the anonymous database while respecting your privacy, guaranteeing continued anonymity and treating all answers confidentially. The researchers only receive and process the information that you have entered on the following pages, and never your name or contact details.

**Contact**

If you have any questions or if any problems arise when completing the survey, you can contact the Helpdesk.

### Part 1: Background questions

Q1: Your gender

- o Female (1)
- o Male (2)

Q2 Your age in years

*Check if number [18-85]*

Q3 Your postal code

Q4 What is your highest completed level of education? *(Single answer)*

o Primary education (1)
o Secondary education – years 7 to 11, age 11 to 16, leading to GCSE (2)
o NVQ levels 1 and 2 (3)
o B-Tech (4)
o A Levels (5)
o Higher education – Bachelor's / Master's degree (6)
o Higher education - PhD (7)
o Other, namely: (8)

Q5 Which of the situations below best describes your current family situation *(Single answer)*

O Single (1)
o Single parent, living (part or full time) with children (2)
o Living with a partner (married or not) and without children (3)
o Living with a partner (married or not) and with children (4)
o None of the above (5)

If Q5= (2) or (4) show Q6

Q6 How many children aged under 18 years do you have? *(Single answer)*

o 0 (1)
o 1 (2)
o 2 (3)
o 3 (4)
o 3 + (5)

If Q6 is shown and if Q6≠(1) show Q7

Q7 In which year was your youngest child born?

*Open field. + check whether 4 digits + check if <=2018 + check if >= 1999*

Q8 What is your mother's country of birth?

o United Kingdom (1)
o Another country in Western Europe (2)
o Another, non-Western European country (3)

Q9 Do you work in or have you ever worked in health care?

o No (1)
o Yes. What was your position? (2)
o Nurse (2.1)
o Doctor (2.2)
o Paramedic (2.3)
o Pharmacist (2.4)
o Other (2.5)

Q10 Have you ever had to deal with a serious illness yourself? (For example cancer)

o Yes (1)
o No (2)

Q11 Do you have the annual flu jab?

o Never (1)
o Sometimes (2)
o Mostly (3)
o Always (4)

Q12 Do you smoke or have you ever smoked?

o Yes, I smoke (1)
o I have smoked, but have stopped (2)
o I have never smoked (3)

Q13 Do any of the convictions below influence the way you think about vaccination? Multiple answers possible. *If 1 answer is chosen, cannot pick 5 Multiple answers possible.*

o Religious belief (1)
o Homeopathy (2)
o Naturopathy (3)
o Anthroposophy (4)
o None of the above answers (5)
o Other, namely: (6)

Q14 What faith or belief do you identify with?

o Protestant (Reformed, Church of England, etc.) (1)
o Catholic (2)

o Islam (3)
o Judaism (4)
o Buddhism or Hinduism (5)
o Other religion or belief (6)
o No religion (7)
o I prefer not to answer this question (8)

If Q6 >1 show Q15

Q15 Have you had your child(ren) vaccinated according to the National Immunization schedule?

o Yes (1)
o No (2)
o Partially (3)
o Don't know (3)
o Not applicable (4)

If Q15 == 1 or 3 show Q17

Q16 What has your experience been of having your children vaccinated ?

o Good (1)
o Bad (2)
o Not applicable (3)

*From now, until the end of the questionnaire, questions are either for group A, for group B or for all participants. Questions for all participants are displayed as Qx. Group A consists of respondents who fill in the questionnaire for themselves. There are no explicit restrictions for this group. So respondents that apply for group B too can, by randomisation, be assigned to group A. Questions exclusively for group A are displayed as QxA*
*Groep B consists only of respondents with at least one child <18y Check by Q6. If Q6 is shown and ≠ (1), than assign persons to group B. This group fills in the questionnaire for decisions concerning*
*their youngest child. Questions exclusively for group B are displayed as QxB.*

Q17B Below are a number of questions with statements about the vaccination of children. These statements concern decisions **about the vaccination of your youngest child.**
Please choose the option that best describes your opinion for every statement *Single answer for each statement*

| | | | | | |
|---|---|---|---|---|---|
| I think that vaccinating my child according to the National Vaccination Program is… | Very unwise | Unwise | Neutral | Wise | Very wise |

| | | | | | |
|---|---|---|---|---|---|
| I think that vaccinating my child according to the National Vaccination Program is… | Very unimportant | Unimportant | Neutral | Important | Very important |
| I think that vaccinating my child according to the National Vaccination Program is… | Very unnecessary | Unnecessary | Neutral | Necessary | Very necessary |
| The people who are important to me think that I must have my child(ren) vaccinated. | Completely disagree | Disagree | Neutral | Agree | Completely agree |
| The people who are important to me have their child(ren) vaccinated. | Completely disagree | Disagree | Neutral | Agree | Completely agree |
| The people who are important to me are pleased if I have my child vaccinated. | Completely disagree | Disagree | Neutral | Agree | Completely agree |
| If I do not have my child vaccinated, there is a good chance that he/she will get an infectious disease that can be vaccinated against within the National Vaccination Program. | Completely disagree | Disagree | Neutral | Agree | Completely agree |
| The National Vaccination Program is good for protecting my child's health. | Completely disagree | Disagree | Neutral | Agree | Completely agree |
| It is important to me that vaccinating my child contributes to the protection of others. | Completely disagree | Disagree | Neutral | Agree | Completely agree |
| The diseases that are vaccinated against can be very serious. | Completely disagree | Disagree | Neutral | Agree | Completely agree |
| The side effects of the vaccinations within the National Vaccination Program can be very serious. | Completely disagree | Disagree | Neutral | Agree | Completely agree |
| Vaccinating my child is something that I do not have to think about for long. | Completely disagree | Disagree | Neutral | Agree | Completely agree |
| Vaccinating my child is the logical thing to do. | Completely disagree | Disagree | Neutral | Agree | Completely agree |

| | | | | | |
|---|---|---|---|---|---|
| If I had to make the choice now, I would have my child vaccinated within the National Vaccination Program. | Completely disagree | Disagree | Neutral | Agree | Completely agree |
| If I had to make an additional appointment with my care provider (* care provider is your GP or child healthcare professional/paediatrician) for vaccination, that would be a reason for not having my child(ren) vaccinated. | Completely disagree | Disagree | Neutral | Agree | Completely agree |
| I have confidence in the information about vaccinations that my care provider (* care provider is your GP or child healthcare professional/paediatrician) gives me. | Completely disagree | Disagree | Neutral | Agree | Completely agree |
| I have confidence in the information about vaccinations that I receive from the Government. | Completely disagree | Disagree | Neutral | Agree | Completely agree |
| I think that it is the responsibility of every parent to have his/her child vaccinated within the National Vaccination Program . | Completely disagree | Disagree | Neutral | Agree | Completely agree |
| I think it is bad if parents do not have their children vaccinated within the National Vaccination Program. | Completely disagree | Disagree | Neutral | Agree | Completely agree |
| Experiencing infectious diseases contributes to the positive mental and physical development of my children. | Completely disagree | Disagree | Neutral | Agree | Completely agree |
| Experiencing infectious diseases leads to better and longer protection than vaccinations against the same diseases. | Completely disagree | Disagree | Neutral | Agree | Completely agree |

Q17A Below are a number of questions with statements about vaccination. The statements concern the decision **to get vaccinated yourself**. Please choose the option that best describes your opinion for every statement *Single answer for each statement*

| | | | | | |
|---|---|---|---|---|---|
| I think that getting vaccinated against infectious diseases is… | Very unwise | Unwise | Neutral | Wise | Very wise |
| I think that getting vaccinated against infectious diseases is… | Very unimportant | Unimportant | Neutral | Important | Very important |
| I think that getting vaccinated against infectious diseases is… | Very unnecessary | Unnecessary | Neutral | Necessary | Very necessary |
| The people who are important to me think that I must get vaccinated. | Completely disagree | Disagree | Neutral | Agree | Completely agree |
| The people who are important to me get vaccinated. | Completely disagree | Disagree | Neutral | Agree | Completely agree |
| The people who are important to me are pleased if I get vaccinated. | lCompletely disagree | Disagree | Neutral | Agree | Completely agree |
| If I do not get vaccinated, there is a good chance that I will get an infectious disease that can be vaccinated against. | Completely disagree | Disagree | Neutral | Agree | Completely agree |
| The available vaccinations are suited to protect my health. | Completely disagree | Disagree | Neutral | Agree | Completely agree |
| It is important to me that getting vaccinated contributes to the protection of others. | Completely disagree | Disagree | Neutral | Agree | Completely agree |
| The diseases that are vaccinated against can be very serious. | Completely disagree | Disagree | Neutral | Agree | Completely agree |
| The side effects of the vaccinations within the National Vaccination Program can be very serious. | Completely disagree | Disagree | Neutral | Agree | Completely agree |
| Getting vaccinated is something that I do not have to think about for long. | Completely disagree | Disagree | Neutral | Agree | Completely agree |
| Getting vaccinated is the logical thing to do. | Completely disagree | Disagree | Neutral | Agree | Completely agree |

| | | | | | |
|---|---|---|---|---|---|
| If I had to make the choice now, I would get vaccinated. | Completely disagree | Disagree | Neutral | Agree | Completely agree |
| If I had to make an additional appointment with my care provider (* care provider is your GP or child healthcare professional/paediatrician) for vaccination, that would be a reason for not getting vaccinated. | Completely disagree | Disagree | Neutral | Agree | Completely agree |
| I have confidence in the information about vaccinations that my care provider (* care provider is your GP or child healthcare professional/paediatrician) gives me. | Completely disagree | Disagree | Neutral | Agree | Completely agree |
| I have confidence in the information about vaccinations that I receive from the Government. | Completely disagree | Disagree | Neutral | Agree | Completely agree |
| I think that it is everyone's responsibility to get vaccinated. | Completely disagree | Disagree | Neutral | Agree | Completely agree |
| I think it is bad if people do not get vaccinated within the National Vaccination Program. | Completely disagree | Disagree | Neutral | Agree | Completely agree |
| Experiencing infectious diseases contributes to positive mental and physical development. | Completely disagree | Disagree | Neutral | Agree | Completely agree |
| Experiencing infectious diseases leads to better and longer protection than vaccinations against the same diseases. | Completely disagree | Disagree | Neutral | Agree | Completely agree |

Q18B Which people are important to you if you have to decide whether or not to have your child vaccinated? (Multiple answers possible)

*Randomize order of choices, 8 always last. Multiple answers*

o Partner (1)
o Family (2)

o Friends (3)
o Other parents (4)
o Colleagues (5)
o GP (6)
o The National Health Service (NHS) (7)
o Other, namely: (8)

Q18A Which people are important to you if you have to decide whether or not to get vaccinated? (Multiple answers possible)

*Randomize order of choices, 8 always last. Multiple answer possible*

o Partner (1)
o Family (2)
o Friends (3)
o Colleagues (4)
o GP (5)
o The National Health Service (NHS) (6)
o Other, namely: (7)

Q19 The following questions describe two situations where you can make a choice *Descriptive text.*

Q20A We provide a simplified example below. Each time, we ask in which of the two situations **you would be most likely to have a vaccine administered**. Some of the features of the two options are different and are highlighted in yellow. *Single answer*

| | |
|---|---|
| 30% of the British population has already been vaccinated | 30% of the British population has already been vaccinated |
| Mild side effects are rare and serious side effects are very exceptional | Mild side effects are common and serious side effects are very exceptional |
| O | o |

Q20B We provide a simplified example below. Each time, we ask in which of the two situations **you would be most likely to have a vaccine administered to your child**. Some of the features of the two options are different and are highlighted in yellow. *Single answer*

| | |
|---|---|
| 30% of the British population has already been vaccinated | 30% of the British population has already been vaccinated |
| Mild side effects are rare and serious side effects are very exceptional | Mild side effects are common and serious side effects are very exceptional |
| O | o |

Q21 --- *10 choice sets from conjoint design: excel file* ---

Q22A <u>How serious</u> would you estimate the impact on your health to be if you get one of the following diseases in the next year? *Single answer for each indication*

| **Disease severity** | Very serious (1) | Serious (2) | Moderately serious (3) | Not serious (4) | Not at all serious (5) |
|---|---|---|---|---|---|
| Flu/influenza (1) | o | o | o | o | o |
| Leukaemia (blood cancer) (2) | o | o | o | o | o |
| Bladder infection (3) | o | o | o | o | o |
| Measles (4) | o | o | o | o | o |

Q22B <u>How serious</u> would you estimate the impact on your youngest child's health to be if your child gets one of the following diseases in the next year? *Single answer for each indication*

| **Disease severity** | Very serious (1) | Serious (2) | Moderately serious (3) | Not serious (4) | Not serious at all (5) |
|---|---|---|---|---|---|
| Flu/influenza (1) | o | o | o | o | o |
| Leukaemia (blood cancer) (2) | o | o | o | o | o |
| Bladder infection (3) | o | o | o | o | o |
| Measles (4) | o | o | o | o | o |

Q23A How high do you estimate <u>the risk</u> that you will get one of the following diseases during the next 12 months. *Single answer for each indication*

| **Risk of diseases** | Very big risk (1) | Big risk (2) | Average risk (3) | Small risk (4) | Very small risk (5) |
|---|---|---|---|---|---|
| Flu/influenza (1) | o | o | o | o | o |
| Leukaemia (blood cancer) (2) | o | o | o | o | o |
| Bladder infection (3) | o | o | o | o | o |
| Measles (4) | o | o | o | o | o |

Q23B How high do you estimate <u>the risk</u> that your youngest child will get one of the following diseases during the next 12 months. *Single answer for each indication*

| **Risk of diseases** | Very big risk (1) | Big risk (2) | Average risk (3) | Small risk (4) | Very small risk (5) |
|---|---|---|---|---|---|
| Flu/influenza (1) | o | o | o | o | o |
| Leukemia (blood cancer) (2) | o | o | o | o | o |
| Bladder infection (3) | o | o | o | o | o |

| | | | | | |
|---|---|---|---|---|---|
| Measles (4) | o | o | o | o | o |

Q24 How do you get or search for information about infectious diseases and vaccination? Multiple answers possible. *Randomize order of choices. Social media never first. Multiple answers possible*

o via social media (Facebook, Twitter, etc.) (1)
o via traditional media (newspaper, TV or magazine) (2)
o via de website of the NHS, or Public Health England (3)
o via the Internet (other than social media) (4)
o via friends and/or family (5)
o via my GP, child healthcare professional or paediatrician (6)
o via my pharmacist (7)
o None of the above (8)

Q25 The information you receive about your health or diseases in general can be unclear or too complicated. We would like to know about your experiences in this regard. *Descriptive text*

Q26 How often does someone help you to read letters or brochures from your GP or the hospital?

o Never (1)
o Occasionally (2)
o Sometimes(3)
o Frequently (4)
o Always (5)

Q27 How confident are you that you can complete medical forms properly yourself?

o Very confident (1)
o Quite confident (2)
o Somewhat confident (3)
o Not very confident (4)
o Not confident at all (5)

Q28 How often is it difficult for you to learn more about your health because you do not understand written information very well?

o Never (1)
o Occasionally (2)
o Sometimes(3)
o Frequently (4)
o Always (5)

Q29 Thank you for participating in our survey. *Descriptive text*
